# Ethnic differences in early onset multimorbidity and associations with health service use, long-term prescribing, years of life lost, and mortality: an observational study using person-level clustering in the UK Clinical Practice Research Datalink

**DOI:** 10.1101/2023.03.03.23286751

**Authors:** Fabiola Eto, Miriam Samuel, Rafael Henkin, Meera Mahesh, Tahania Ahmad, Alisha Angdembe, R. Hamish McAllister-Williams, Paolo Missier, Nick J Reynolds, Michael R Barnes, Sally Hull, Sarah Finer, Rohini Mathur

**Author notes:** These authors contributed equally (Sarah Finer, Rohini Mathur).

## Abstract

**Background:** The population prevalence of multimorbidity (the existence of at least 2 or more long-term conditions (LTCs) in an individual) is increasing among young adults, particularly in minority ethnic groups and individuals living in socioeconomically deprived areas. In this study, we applied a data-driven approach to identify clusters of individuals who had an early onset multimorbidity in an ethnically and socioeconomically diverse population. We identified associations between clusters and a range of health outcomes.

**Methods and findings:** We analysed the electronic health records from 837,869 individuals in England with early onset multimorbidity (aged between 16 and 39 years old when the second LTC was recorded) using linked primary and secondary care data between 2010 and 2020 from the Clinical Practice Research Datalink GOLD (CPRD GOLD). A total of 204 LTCs were included. Latent class analysis stratified by ethnicity unveiled 4 clusters of multimorbidity in White groups and 3 clusters in South Asian and Black groups. We found that early onset multimorbidity is the most common form of multimorbidity among minority ethnic (59% and 56%, in the South Asian and Black populations, respectively) in the UK compared to the White population (42%). At the end of the study, 4% of the White early onset multimorbidity population had died compared to 2% of the South Asian and Black populations, however, the latter groups died younger and lost more years of life. The three ethnic groups displayed a cluster of individuals with increased rates of primary care consultations, hospitalisations, long-term prescribing, and odds of mortality. These presented a combination of physical and mental health conditions that are common across all groups (hypertension, depression and painful conditions being the leading conditions). However, they also presented exclusive LTCs and had different sociodemographic profiles: Whites were mostly men (54%), South Asian and Black groups were more socioeconomically deprived than White groups, with a consistent deprivation gradient across all multimorbidity clusters. In White groups, the highest risk cluster was more socioeconomically deprived than the lowest risk cluster.

**Conclusions:** These findings emphasise the need to identify, prevent and manage multimorbidity early in the life course. Our work provides additional insights into the need to ensure healthcare improvements are equitable and reach those from socioeconomically deprived and diverse groups who are disproportionately and more severely affected by multimorbidity.

## Introduction

The growing prevalence of multimorbidity – the existence of multiple long-term conditions (LTCs) in a single individual [1]– and its burden on individual and population health has recently led to major research investment and health policy initiatives [2,3].

By 2035, two-thirds of adults over 65 years old in the UK are expected to have multiple long-term conditions [4], and similar trends are observed globally [1]. These trends are coupled with a decrease in the age of onset of multimorbidity [4,5], highlighting the need for effective public health policies to identify, risk stratify and prevent their onset early in the life course.

Although many studies of multimorbidity focus on older adults since its prevalence increases with ageing [4,6], few have investigated multimorbidity in younger populations [5,7]. Importantly, recent studies have shown an increasing prevalence of multimorbidity in early adulthood [4,8]. Similarly, there is evidence showing that individuals with socioeconomic vulnerability experience poorer health outcomes, such as lower quality healthcare provision, premature death and higher mortality rates. Likewise, some minority ethnic groups and individuals living in deprived areas have their onset of multimorbidity earlier in life [8–10].

People living with multiple LTCs account for the majority of primary care and hospital utilisation and long-term medication use. Systematic reviews on the impact of multimorbidity on healthcare utilisation and costs have shown that in the UK [11] and in different countries [12] health service utilisation and costs tend to increase with each additional condition in a single individual. Nonetheless, medical guidelines and strategies to improve health care are centred on the treatment of individual health conditions and very often do not account for the management of multiple and complex conditions that commonly co-occur [13,14]. Likewise, each additional condition has been associated with an increased mortality risk and even higher risk in some ethnic minority groups (Pakistani, Black African, Black Caribbean and Other Black ethnic groups) [10].

A comprehensive approach to map patterns of multiple LTCs in an ethnically and socioeconomically diverse population with early onset of multimorbidity is crucial to understand the distinct and shared mechanisms that lead to disease accumulation and enable early intervention and reconfigure services to meet the needs of more vulnerable groups.

The majority of studies on multimorbidity focus on diseases rather than individuals to assess multimorbidity patterns [15]. However, analysing multimorbidity patterns at an individual level enables a deeper understanding of potentially shared biological and environmental risk factors among specific population groups and understand what similarities they share in terms of sociodemographic profile and what LTCs are the main drivers of increased healthcare service utilisation, long term prescribing and mortality.

Most definitions of multimorbidity state that it is the presence of two or more long-term conditions, yet the majority of multimorbidity research in the UK focuses on a limited set of around 40 highly prevalent long-term conditions [16–21]. This limited set of LTCs leads to the exclusion of less prevalent or ethnically patterned diseases that may have major impact to individuals and health systems. Additionally, the selection of limited numbers of long-term conditions in case definitions is likely to lead to substantial underestimation of population prevalence of multimorbidity. A recent systematic review of multimorbidity studies also highlights the variable and poorly reported measurements of multimorbidity and suggests the need for consensus-based, reproducible definitions [22]. Building on these limitations of existing multimorbidity research, and based on published sources [23–26] we developed and tested a consensus-derived, open-access codelist resource [27] for multimorbidity research with an expanded focus on all LTCs that might contribute to multimorbidity, irrespective of prevalence, with a particular ambition that this resource can adequately address ethnic differences in multimorbidity presentations that may be driven by low prevalence but high impact conditions that are ethnically patterned.

Using electronic health records derived from large national databases we applied a data-driven approach – which is more likely to reveal underlying disease relationships within clusters – to identify patterns of long-term conditions that are more prevalent in clusters of individuals who had an early onset of multimorbidity in an ethnically and socioeconomically diverse multimorbid population. Additionally, we assessed whether the clusters of individuals across ethnic groups have different associations with clinically meaningful health outcomes: health service utilisation, long-term prescribing, years of life lost and mortality.

## Methods

### Study Population and Data Source

We performed a cross-sectional study using electronic health records (EHR) from the Clinical Practice Research Datalink GOLD (CPRD GOLD), a large representative English dataset. We assessed data from individuals aged 16 years and over, permanently registered in any English General Practice between January 1, 2010, and December 31, 2020. All individuals who met CPRD’s acceptable data quality criteria for use in research and who had linkage data available from the Hospital Episode Statistics (HES) for admitted-patient care were selected to be included in our study. We also obtained linkage to national death registries (Office for National Statistics) and the socioeconomic deprivation data (Index of Multiple Deprivation - IMD) for individuals in the source population.

From the CPRD GOLD source population, we selected a study population of individuals who met the following inclusion criteria: (i) had at least two out of a list of 204 long-term conditions, (ii) belong to one of the following ethnic groups: White, South Asian or Black or Black British, (iii) have the date when the LTC was recorded in order to calculate the individual’s age at the onset of multimorbidity, and (iv) have had an early onset of multimorbidity, defined by having the second LTC recorded between the ages of 16 and 39. The selection of our source population is illustrated in the Supplementary file, section 2.

### Defining multimorbidity

We used the definition of multimorbidity set out by the Academy of Medical Sciences [1], as follows: The co-existence of two or more long-term conditions, each one of which is either (a) a physical non-communicable disease of long duration, such as cardiovascular disease or cancer, (b) a mental health condition of long duration, such as a mood disorder or dementia, (c) an infectious disease of long duration, such as HIV or hepatitis C.

Building on existing literature and previous concerns about the lack of reproducibility in multimorbidity research, we undertook a systematic approach to operationalising this definition of multimorbidity for our study. We searched the literature for definitions of multimorbidity and made comparisons between LTCs included in different studies [16,17,23–25]. We searched existing online repositories, publications and supplementary material for previously-built codelists. Where multiple codelists were found, we combined all the relevant codes used by the studies to develop a baseline codelist that underwent extensive clinical revision. We established a clinical consensus exercise to further review the LTCs identified in the studies selected and to assess whether each condition met the definition of multimorbidity we adopted.

Following a two-round consensus exercise composed of ten clinicians from diverse specialities, 204 LTCs were included in our operational definition of multimorbidity from a base list of 311 LTCs, derived from existing high quality studies and codelist resources studies [16,17,23–25]. The codelist revision, development and curation consisted of the following approaches – (a). Comparison between existing codelists followed by clinical curation, (b). Development of new codelists through relevant medical terms and, (c). Cross-mapping of different code systems using revised codelists as a baseline. We revised 263 codelists built on Read v2, 214 codelists built on ICD-10 (International Classification of Diseases, Tenth Revision), 13 codelists built on OPCS-4 (Classification of Interventions and Procedures version 4), and 2 codelists built on gemscript codes (prescribing codes). Detailed information on the methods used to curate the codelists, and the codelists themselves, are available in the MULTIPLY-Initiative online repository [27].

We considered prevalent cases of the LTCs as the presence of any relevant code ever recorded in the individual’s EHR and defined age of onset as the first occurrence of a relevant clinical code. Therefore, by definition, if a condition was never recorded it was considered absent.

### Stratification groups

To investigate whether the accumulation of LTCs over the life course is ethnically patterned in people with early onset of multimorbidity we stratified our analysis according to three following ethnic groups – White, Black or Black British (Black African, Black Caribbean and Black Other) and South Asian (Indians, Bangladeshis and Pakistanis). Information on ethnicity was obtained from self-reported information held in electronic health records and captured during primary care registration and/or consultation episodes [28].

### Covariables

We described the characteristics of people in our population at the age at the start of the study (2010), age at onset of multimorbidity, sex and socioeconomic deprivation (index of multiple deprivation in quintiles, where the 1st quintile represents the least deprived areas and the 5th quintile, the most deprived).

### Defining outcomes

We selected the following adverse outcomes with major impact to people living with multimorbidity and healthcare systems: health service utilisation, long-term prescribing, years of potential life lost (YLL), and mortality. Health service utilisation was defined as the number of primary care consultations (defined by the dates of consultation with any primary care clinician and regardless of the type of consultation), and the number of hospitalisations (defined by the discharge dates related to admitted-patient care) recorded between 2010 and 2020. Long-term prescribing was identified as the counts of unique prescriptions per BNF (British National Formulary) subparagraphs [29], prescribed 3 or more times per year and it was assessed for the 10-year study period (2010 to 2020). Odds of mortality were assessed at the years 5^th^ and 10^th^ after the start of the study and were based on the total number of deaths by the end of the 5^th^ and 10^th^ year after 2010, respectively. The YLL for each ethnic group and their respective clusters of individuals were estimated using the R library ‘*lillies*’ [30] which allows the estimation of YLL according to a given condition (e.g. groups of individuals with a certain characteristic), and the calculation of confidence intervals using bootstrapping technique. The years of life lost were estimated using the remaining average life expectancy after becoming multimorbid and before reaching the average life expectancy at birth of 81 years for the United Kingdom population [31].

### Statistical analysis

#### The sample characteristics were described by ethnic group

To focus on people rather than diseases as the unit of analysis, we applied the latent class analysis (LCA) to unveil different profiles of people who shared similar patterns of LTCs accumulation throughout life. This approach allows each LTC to appear in multiple subgroups of individuals and it is more consistent with clinical experience than other approaches where each LTC could belong to only one cluster at a time. LCA is a person-centred mixture modelling that identifies latent or unobserved classes (e.g. subpopulations) within a sample based on their patterns of responses to observed variables (e.g. presence/absence of an LTC) given by the posterior membership probabilities which inform the probability of an individual belonging to a certain subgroup [32]. For each latent class (e.g. subgroup of people with similar characteristics), the average latent class probability is estimated, which indicates the probability of the class model accurately predicting class membership for individuals [33].

For each ethnic group, we tested 2 to 10-class models (where the number of classes represents the number of possible clusters) with a maximum iteration of 1,000 using the *poLCAParallel* [34] R package and R-4.2.1. We obtained and compared the fit statistics for each *k*-class model, which along with clinical judgement – on the clusters that besides being clinically meaningful also captured heterogeneous LTCs – supported the selection of the optimal number of latent classes. We obtained the bootstrapped likelihood ratio test (BLRT) which indicates if a model with *k* classes is statistically better than a model with *k–1* class upon a provided *p-value* [35]. We evaluated the Bayesian Information Criterion (BIC), the Sample-size Adjusted Bayesian Information Criterion (SABIC), and the likelihood ratio test (LRT) to compare competing models. They inform how much a model is improved by an additional class. Lower BIC, SABIC and LRT values indicate a better fit model. The entropy was assessed to verify the model quality since it indicates how accurately the model defines classes. In general, a value closer to 1 is desirable, although there is no agreed-upon cutoff criterion for entropy [36]. More information on the model selection criteria can be read in the Supplementary file section 3.

We described the distribution of the LTCs per cluster as well as the characterisation of each cluster of people according to sociodemographic variables, health service utilisation, years of life lost and mortality. Generalised linear models adjusted by age at the study start, sex and deprivation level were estimated to investigate which clusters had higher odds of mortality, greater years of life lost (YLL), and higher health services utilisation over a 10 years interval. Logistic regression models were fitted to investigate cluster differences in the odds of mortality by the end of the 5^th^ and 10^th^ years. To account for the overdispersion found in the number of consultations and hospitalisations we fitted negative binomial regression models and zero-inflated Poisson models to deal with the excess of zeros found in long-term prescribing data. For each outcome of interest, the cluster with the smallest association was considered the reference group.

## Results

### Description of the multimorbid study population

Using the consensus exercise outlined, we extensively revised and selected codelists for 204 LTCs defined by 11,053 Read v2 codes, 2,594 ICD-10 codes, 747 OPCS-4 codes, and 3,829 gemscript codes. Detailed information on the codelist curation and development and all codelists used in our study can be found in the MULTIPLY-Initiative online repository [27].

A total of 2,814,507 individuals had at least one of the 204 LTCs ever recorded in their EHR from the source English population. We then identified our multimorbid population (n = 1,961,888 (69.7%) as those who had developed two or more of the 204 LTCs at age 16 years or above, and who belonged to one of the three ethnic groups under investigation (Supplementary file section 2).

### Description of the early onset multimorbid population

From the total multimorbid population, we identified 837,869 individuals with early onset multimorbidity (16 to 39 years at onset), of whom 777,906 (93%) were White, 33,915 (4%) South Asian and 26,048 (3%) were Black or Black British. Early onset multimorbidity was the most common form of multimorbidity among South Asian and Black groups (59% and 56%, respectively) in contrast with the White population (42%). The median age at multimorbidity onset was 30 years for South Asians, 31 years for the Black and 29 years in the White population. Women comprised the majority of multimorbid individuals, particularly in the South Asian and Black populations (70% and 73% respectively, compared to 65% in the White population). The early onset multimorbid South Asian and Black populations were mostly from greater socioeconomically deprived areas (28% and 39%, respectively, belonged to the most deprived IMD quintile) compared to the White population where 21% belonged to the least deprived IMD quintile. The median number of multiple LTCs was higher in the White population (median = 6, IQR 4-10), compared to the South Asian (5, 3-8) and the Black population (5, 3-8). At the end of the study, 4% (N= 36,027) of the total young multimorbid population died, 4% of Whites, 2% of South Asians and 2% of Blacks. However, South Asians and Blacks died younger than Whites (median age = 52, 48 and 61, respectively) (Table 1) (Supplementary file: Table S1).

**Table 1.** Sociodemographic characteristics of the population with early onset of multimorbidity according to ethnic group. CPRD GOLD (2010-2020).

| Ethnic groups | White | South Asian | Black or Black British | Total |
| --- | --- | --- | --- | --- |
| **Total** | 777,906 (93) | 33,915 (4) | 26,048 (3) | 837,869 (100) |
| **Sex** |  |  |  |  |
| Men | 269,668 (35) | 10,263 (30) | 6,910 (27) | 286,841 (34) |
| Women | 508,238 (65) | 23,652 (70) | 19,138 (73) | 551,028 (66) |
| **Age at onset<sup>1</sup>** |  |  |  |  |
| Median (Q1– Q3) | 29 (23–34) | 30 (25–34) | 31 (25–35) | 29 (23–34) |
| **Age at the study start (2010)** |  |  |  |  |
| Median (Q1- Q3) | 37 (28–46) | 32 (27–39) | 33 (27–40) | 36 (28–45) |
| **Age at death** |  |  |  |  |
| Median (Q1- Q3) | 61 (49–76) | 52 (43–65) | 48 (41–58) | 61 (49–75) |
| **Index Multiple Deprivation (quintiles) (%)** |  |  |  |  |
| 1Q (least deprived) | 1,65,180 (21) | 5,343 (16) | 1,614 (6) | 172,137 (21) |
| 2Q | 159,959 (21) | 5,615 (17) | 2,413 (9) | 167,987 (20) |
| 3Q | 159,372 (21) | 6,301 (19) | 4,366 (17) | 170,039 (20) |
| 4Q | 150,768 (19) | 7,173 (21) | 7,418 (29) | 165,359 (20) |
| 5Q (greatest deprived) | 142,020 (18) | 9,466 (28) | 10,213 (39) | 161,699 (19) |
| **Consultation in 10 years** |  |  |  |  |
| Median (Q1– Q3) | 75 (36–142) | 80 (42–143) | 71 (36–127) | 75 (36–141) |
| **Hospitalisation in 10 years<sup>3</sup>** |  |  |  |  |
| Median (Q1– Q3) | 2 (1–5) | 2 (1–5) | 3 (1–5) | 2 (1–5) |
| **Long-term prescribing in 10 years<sup>4</sup>** |  |  |  |  |
| Median (Q1– Q3) | 2 (0–5) | 2 (0–5) | 1 (0–4) | 2 (0–5) |
| **Mortality** |  |  |  |  |
| 5-year mortality | 12,906 (2) | 194 (1) | 204 (1) | 13,304 (2) |
| 10-year mortality | 34,922 (4) | 535 (2) | 570 (2) | 36,027 (4) |
| **Years of Life Lost<sup>5</sup>** |  |  |  |  |
| Estimate (95% CI) | 21.1 (21.0–21.2) | 28.0 (27.0–29.2) | 30.8 (29.9–31.9) | 21.4 (21.2–21.6) |
| **Remaining life expectancy** |  |  |  |  |
| Estimate (95% CI) | 31.0 (30.8–31.1) | 22.0 (21.2–23.3) | 18.1 (16.9–19) | 31.0 (30.5–31.8) |
| **Long-term conditions** |  |  |  |  |
| Median (Q1– Q3) | 6 (4–10) | 5 (3–8) | 5 (3–8) | 6 (4–9) |

<sup>1</sup> Individual's age when the second LTC was recorded.
<sup>2</sup> Individual's age when one of the earliest occur: study end (31-12-2020) or death.
<sup>3</sup> It does not include Accident and Emergency attendances.
<sup>4</sup> Counts of unique BNF subparagraphs from which an individual had continuous prescribing (3 or more prescriptions that occur in a year).
<sup>5</sup> UK's life expectancy at birth of 81 years old taken as reference.

After evaluating the fit statistics for the latent class models (Supplementary file: section 3, Figures 1 and 2, and Table S3) and upon clinical judgement, we identified four clusters of individuals in the White population and three clusters of individuals in the South Asian and Black populations. Although we included all 204 LTCs in the latent class models, throughout the paper we discuss only the twenty most prevalent conditions in each cluster for clarity. The prevalence for all 204 LTCs per cluster and ethnicity can be found in the Supplementary file: Table S3.

**Figure 1.**
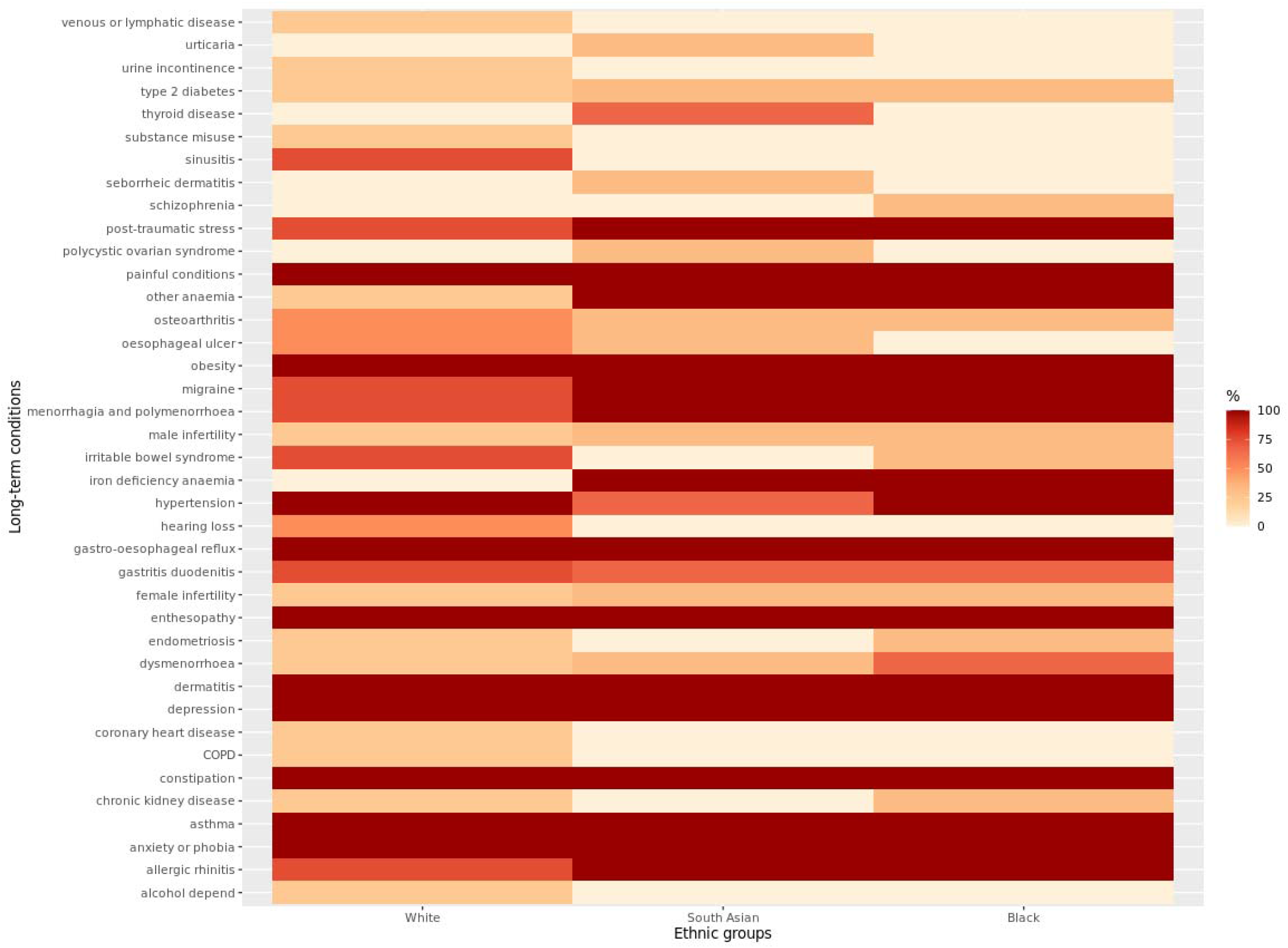
The top 20 most prevalent conditions in the clusters of individuals with early onset of multimorbidity and different ethnic groups. The figure shows the overlapping and unique LTC across ethnic groups and the proportion of clusters they occur within each ethnic group. Maximum number of clusters: White = 4, South Asian = 3, Black = 3.

**Figure 2.**
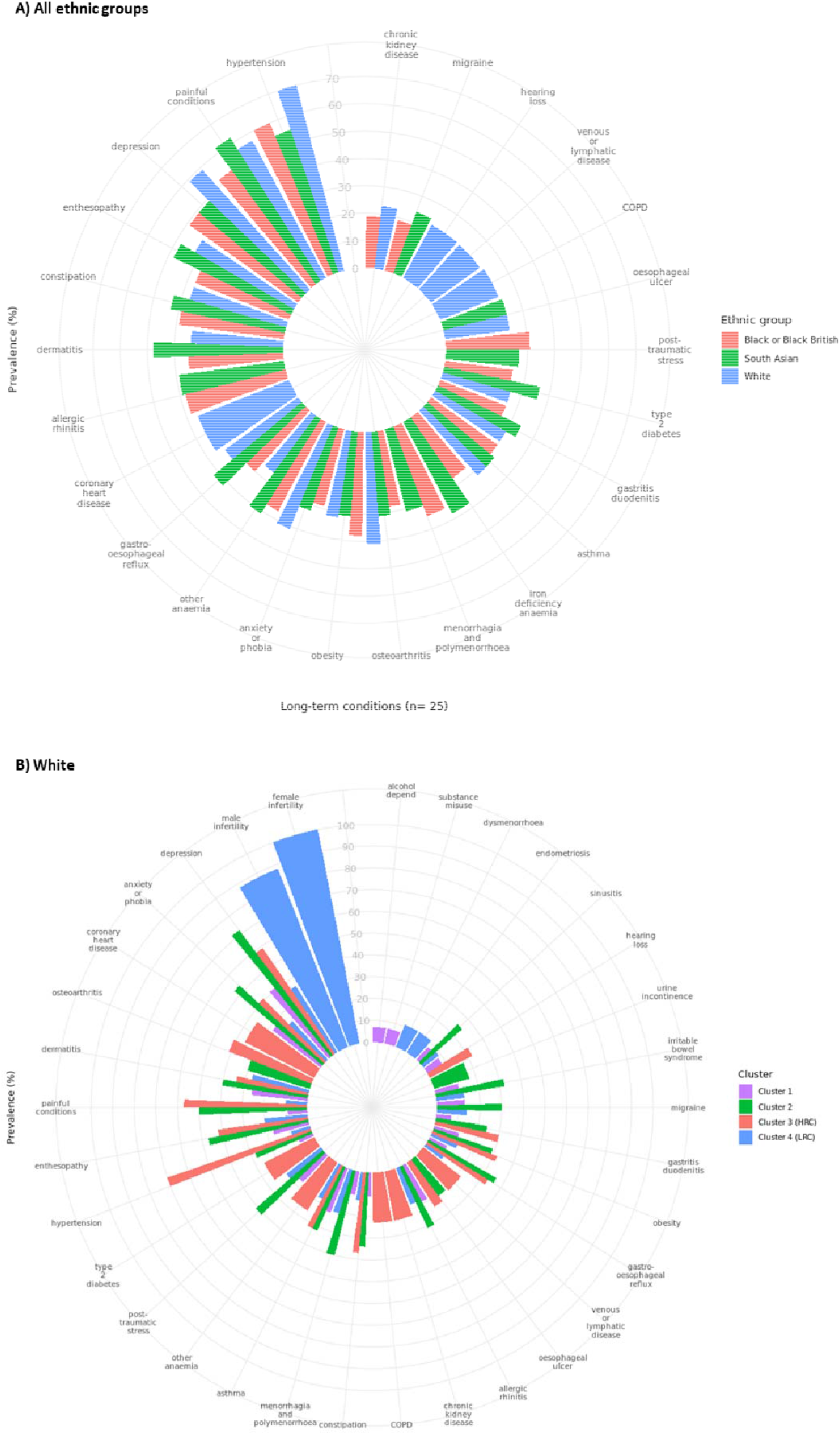
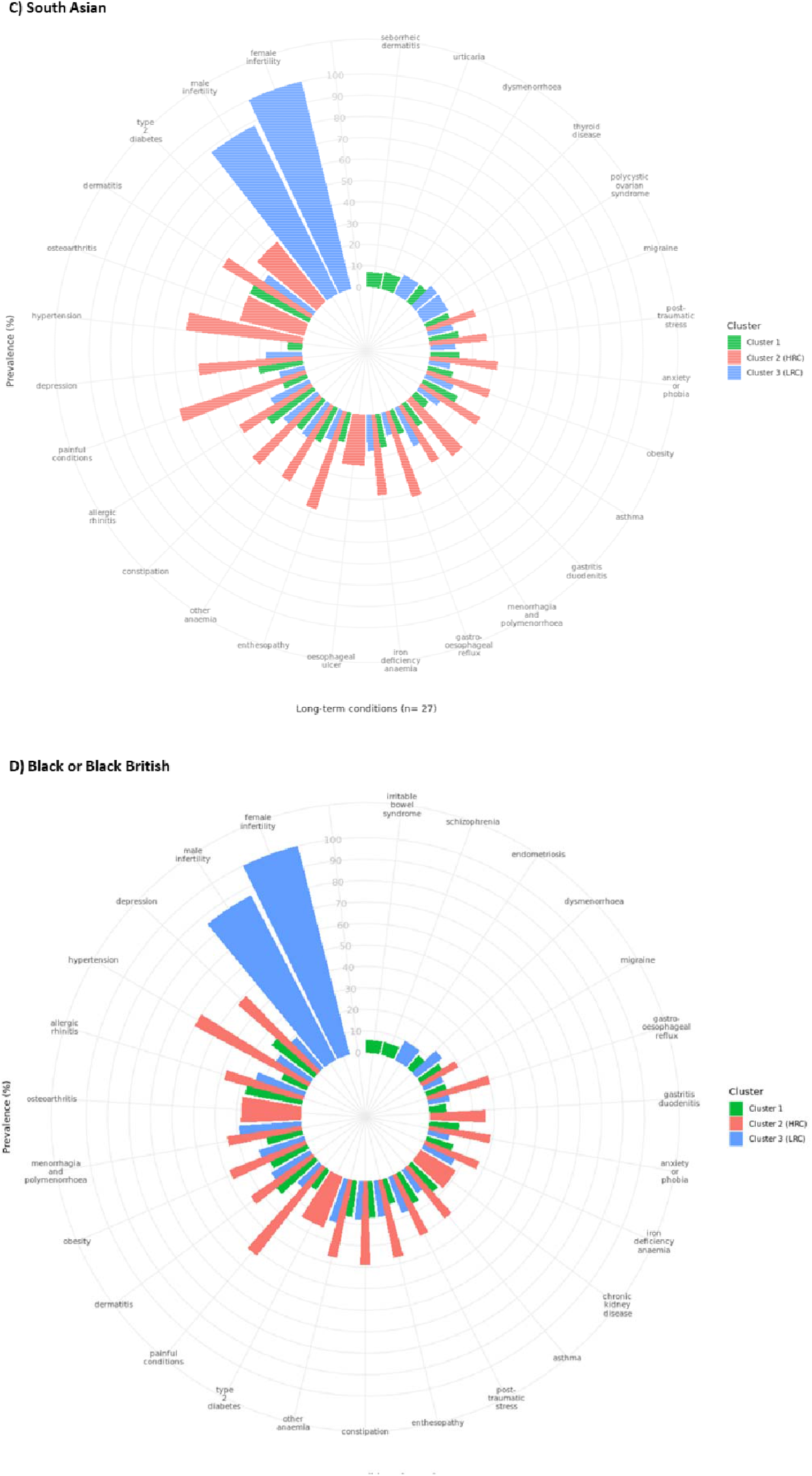
Twenty most prevalent conditions in the clusters (where the difference between within-cluster relative prevalence and prevalence in the ethnic group is the largest). A) Highest-risk clusters of individuals according to ethnic group. B) Clusters of multimorbidity in White individuals. C) Clusters of multimorbidity in South Asian individuals. C) Clusters of multimorbidity in Black or Black British individuals. The plots display common and unique LTCs across clusters of individuals.

Figure 1 shows the distribution of 39 conditions found among the top 20 most prevalent LTCs across all three ethnic groups and the proportion of clusters where each condition appears. Twenty-one LTCs were common among at least one cluster of the three ethnic groups but had varied prevalence. Anxiety or phobia, asthma, constipation, depression, dermatitis, enthesopathy, gastro-oesophageal reflux, obesity and painful conditions were highly prevalent LTCs that occurred in all clusters across the three ethnic groups (Figure 1). There were eight highly prevalent LTCs that appeared exclusively in the White population clusters: alcohol dependence and related disease, chronic obstructive pulmonary disease (COPD), coronary heart disease (CHD), hearing loss, sinusitis, psychoactive substance misuse, urinary incontinence, and venous or lymphatic disease. Four LTCs were identified as highly prevalent in, but exclusive to, the South Asian population clusters, polycystic ovarian syndrome, seborrheic dermatitis, thyroid disease, and urticaria. And finally, in the Black population only, schizophrenia was present as a highly prevalent LTC in clusters of the Black population, but none of the other ethnic groups (Figures 1 and 2).

We next investigated different parameters of healthcare utilisation in this early onset multimorbid population during the 10 year study period (2010-2020). South Asians and Whites had a higher number of primary health care consultations per individual (South Asians median 80, IQR:42-143; Whites 75, 36-142), while Blacks had the lowest number of primary care consultations in the same period (71, 36-127). Similarly, median number of long-term prescriptions between 2010 and 2020 per individual was higher among South Asians (2, 0-5) and Whites (2, 0-5) compared to Blacks (1, 0-4). However, the number of hospitalisation episodes (not including Accident and Emergency attendances) over a 10 year period was higher for Blacks (3,1-5) compared to Whites and South Asians (2, 1-5).

Furthermore, Blacks lose more years of life on average (30.8) than Whites (21.1) and South Asians (28.0) after developing early-onset multimorbidity and before reaching the average UK life expectancy [31] of 81 years old.

### Description and ethnic comparison of high and low risk clusters amongst individuals with early onset multimorbidity

We defined “high risk clusters” (HRC) and “low risk clusters” (LRC) using a composite of primary care consultations, hospitalisation, long-term prescribing and mortality (Table 2 and 3).

**Table 2.**
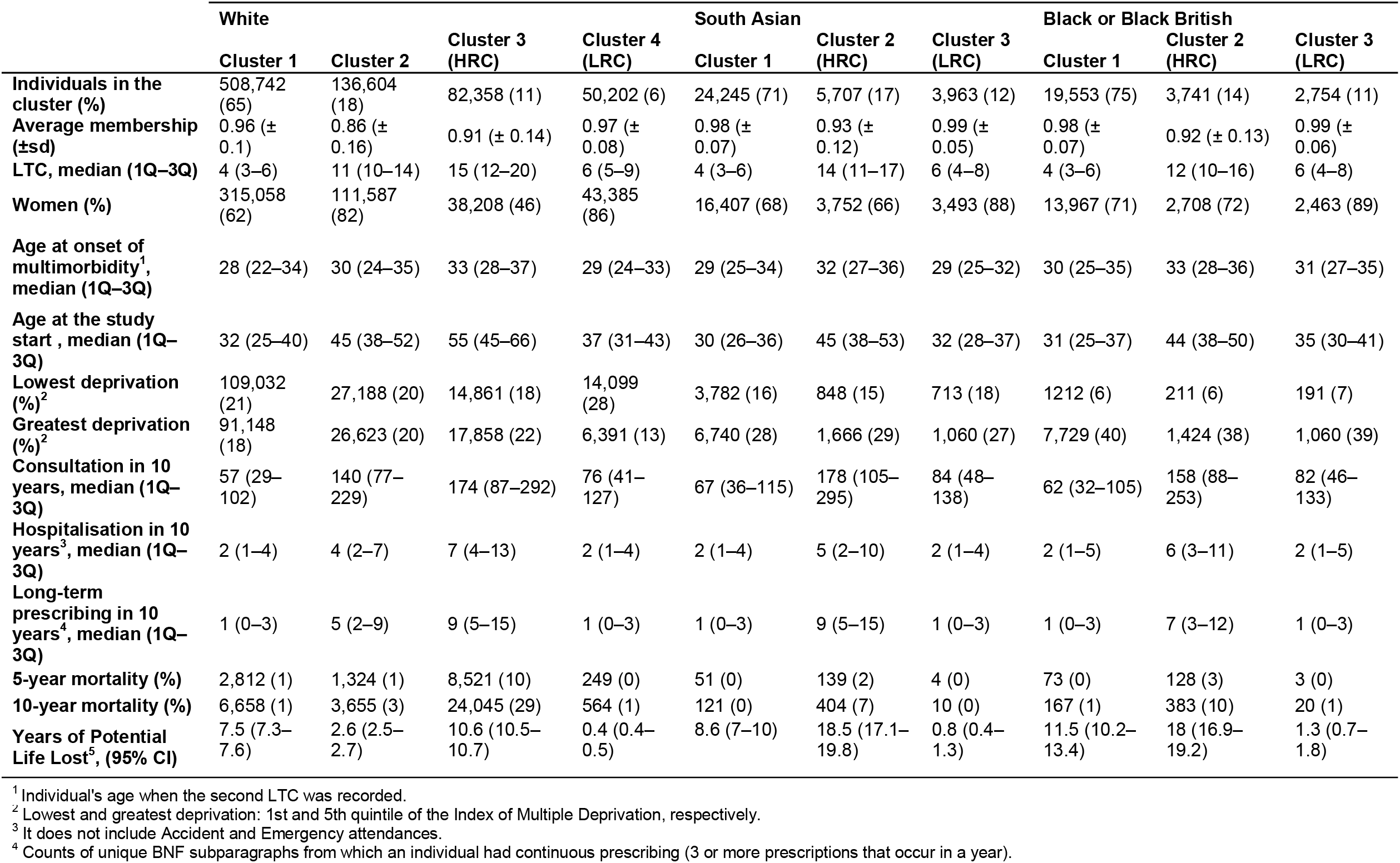
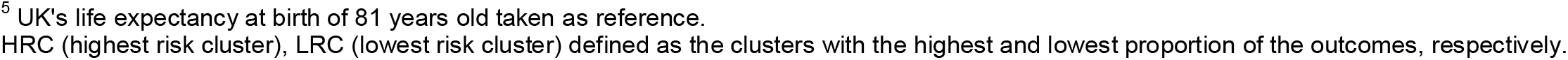
Characteristics of the clusters of individuals with early onset of multimorbidity from different ethnic groups.

|  | White |  |  |  | South Asian |  |  | Black or Black British |  |  |
| --- | --- | --- | --- | --- | --- | --- | --- | --- | --- | --- |
|  | Cluster 1 | Cluster 2 | Cluster 3 (HRC) | Cluster 4 (LRC) | Cluster 1 | Cluster 2 (HRC) | Cluster 3 (LRC) | Cluster 1 | Cluster 2 (HRC) | Cluster 3 (LRC) |
| **Individuals in the cluster (%)** | 508,742 (65) | 136,604 (18) | 82,358 (11) | 50,202 (6) | 24,245 (71) | 5,707 (17) | 3,963 (12) | 19,553 (75) | 3,741 (14) | 2,754 (11) |
| **Average membership (±sd)** | 0.96 (± 0.1) | 0.86 (± 0.16) | 0.91 (± 0.14) | 0.97 (± 0.08) | 0.98 (± 0.07) | 0.93 (± 0.12) | 0.99 (± 0.05) | 0.98 (± 0.07) | 0.92 (± 0.13) | 0.99 (± 0.06) |
| **LTC, median (1Q–3Q)** | 4 (3–6) | 11 (10–14) | 15 (12–20) | 6 (5–9) | 4 (3–6) | 14 (11–17) | 6 (4–8) | 4 (3–6) | 12 (10–16) | 6 (4–8) |
| **Women (%)** | 315,058 (62) | 111,587 (82) | 38,208 (46) | 43,385 (86) | 16,407 (68) | 3,752 (66) | 3,493 (88) | 13,967 (71) | 2,708 (72) | 2,463 (89) |
| **Age at onset of multimorbidity<sup>1</sup>, median (1Q–3Q)** | 28 (22–34) | 30 (24–35) | 33 (28–37) | 29 (24–33) | 29 (25–34) | 32 (27–36) | 29 (25–32) | 30 (25–35) | 33 (28–36) | 31 (27–35) |
| **Age at the study start, median (1Q–3Q)** | 32 (25–40) | 45 (38–52) | 55 (45–66) | 37 (31–43) | 30 (26–36) | 45 (38–53) | 32 (28–37) | 31 (25–37) | 44 (38–50) | 35 (30–41) |
| **Lowest deprivation (%)<sup>2</sup>** | 109,032 (21) | 27,188 (20) | 14,861 (18) | 14,099 (28) | 3,782 (16) | 848 (15) | 713 (18) | 1212 (6) | 211 (6) | 191 (7) |
| **Greatest deprivation (%)<sup>2</sup>** | 91,148 (18) | 26,623 (20) | 17,858 (22) | 6,391 (13) | 6,740 (28) | 1,666 (29) | 1,060 (27) | 7,729 (40) | 1,424 (38) | 1,060 (39) |
| **Consultation in 10 years, median (1Q–3Q)** | 57 (29–102) | 140 (77–229) | 174 (87–292) | 76 (41–127) | 67 (36–115) | 178 (105–295) | 84 (48–138) | 62 (32–105) | 158 (88–253) | 82 (46–133) |
| **Hospitalisation in 10 years<sup>3</sup>, median (1Q–3Q)** | 2 (1–4) | 4 (2–7) | 7 (4–13) | 2 (1–4) | 2 (1–4) | 5 (2–10) | 2 (1–4) | 2 (1–5) | 6 (3–11) | 2 (1–5) |
| **Long-term prescribing in 10 years<sup>4</sup>, median (1Q–3Q)** | 1 (0–3) | 5 (2–9) | 9 (5–15) | 1 (0–3) | 1 (0–3) | 9 (5–15) | 1 (0–3) | 1 (0–3) | 7 (3–12) | 1 (0–3) |
| **5-year mortality (%)** | 2,812 (1) | 1,324 (1) | 8,521 (10) | 249 (0) | 51 (0) | 139 (2) | 4 (0) | 73 (0) | 128 (3) | 3 (0) |
| **10-year mortality (%)** | 6,658 (1) | 3,655 (3) | 24,045 (29) | 564 (1) | 121 (0) | 404 (7) | 10 (0) | 167 (1) | 383 (10) | 20 (1) |
| **Years of Potential Life Lost<sup>5</sup>, (95% CI)** | 7.5 (7.3–7.6) | 2.6 (2.5–2.7) | 10.6 (10.5–10.7) | 0.4 (0.4–0.5) | 8.6 (7–10) | 18.5 (17.1–19.8) | 0.8 (0.4–1.3) | 11.5 (10.2–13.4) | 18 (16.9–19.2) | 1.3 (0.7–1.8) |

<sup>1</sup> Individual's age when the second LTC was recorded.
<sup>2</sup> Lowest and greatest deprivation: 1st and 5th quintile of the Index of Multiple Deprivation, respectively.
<sup>3</sup> It does not include Accident and Emergency attendances.
<sup>4</sup> Counts of unique BNF subparagraphs from which an individual had continuous prescribing (3 or more prescriptions that occur in a year).
<sup>5</sup> UK's life expectancy at birth of 81 years old taken as reference.
HRC (highest risk cluster), LRC (lowest risk cluster) defined as the clusters with the highest and lowest proportion of the outcomes, respectively.

**Table 3.**
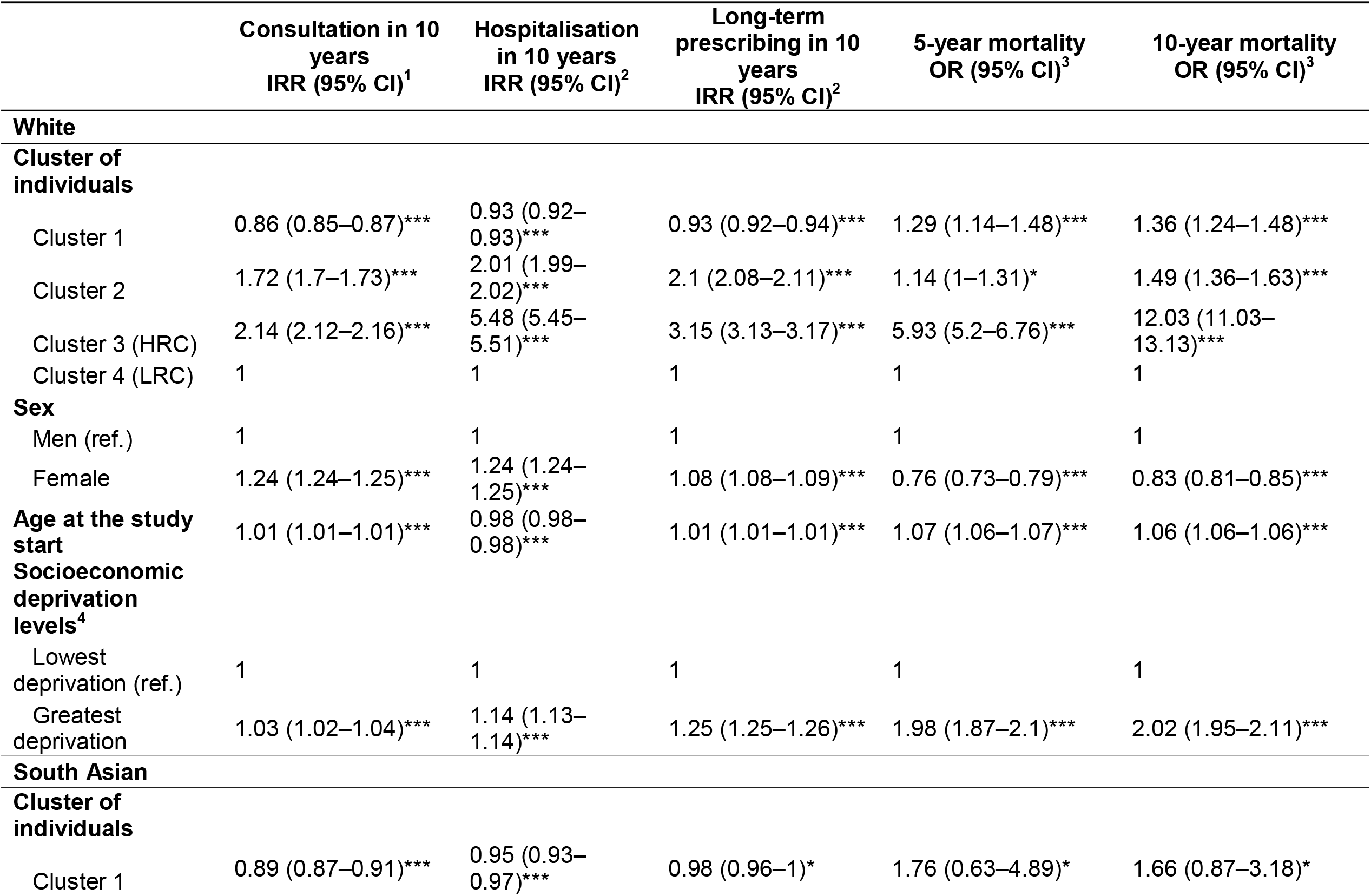
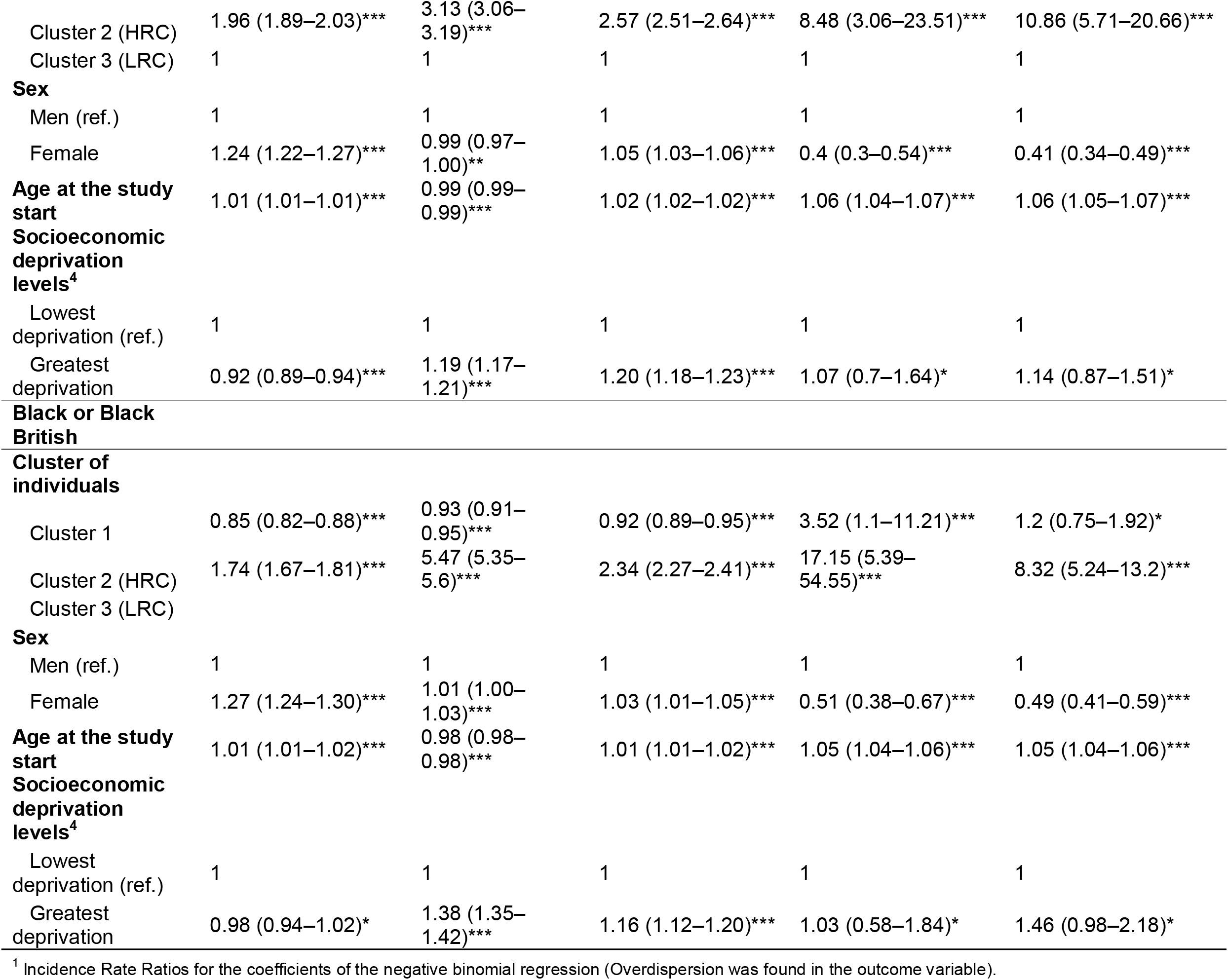
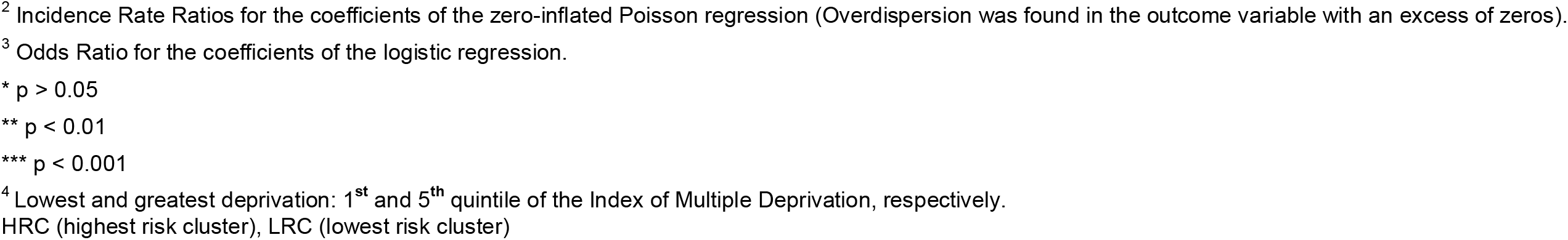
Association between the health service utilisation, mortality and years of life lost and the different clusters of individuals with early onset of multimorbidity according to ethnic groups. All models were adjusted by sex, age at the start of the study and deprivation. The clusters with the lowest impact on the outcomes were considered as references.

The HRCs shared 14 of the 20 most prevalent LTCs and shared their three leading conditions – hypertension, depression and painful conditions (Figure 2A). However, beyond these similarities, there was variation in the composition of HRCs according to ethnicity.

COPD, CHD, hearing loss and venous or lymphatic disease were only found among the most prevalent conditions in Whites. In contrast, allergic rhinitis, iron deficiency anaemia, menorrhagia and polymenorrhoea, migraine, and post-traumatic stress and stress-related disorders (PTSD) were amongst the most prevalent conditions in South Asian and Black groups. Oesophageal ulcer was among the most prevalent conditions in White and South Asian groups, but not in Black groups. Chronic kidney disease was present in Whites and Black groups, but not in South Asians (Figure 2A).

The lowest risk clusters were relatively homogeneous across all 3 ethnic groups and were used as the reference group in our regression models. The most common LTCs in the LRC, across all ethnicities, were female infertility and male infertility. We compared the HRC to the LRC within each ethnic category.

For individuals of White ethnicity, the three most common conditions in HRC were hypertension, depression and painful conditions (Figure 2B), followed by enthesopathy and synovial disorders, osteoarthritis and anxiety or phobias. People in the HRC had a greater median number of LTCs (n=15, IQR 12-20) compared to those in the LRC (6, 5-9) (Table 2); a greater proportion of men (54% in the HRC compared to 14% in the LRC); and a greater numbers of people living in the most deprived IMD quintile (22% vs 13%). Individuals in the HRC had over double the rate of primary care consultation, [IRR= 2.14, 95% CI 2.12–2.16]; three times the rate of long-term prescribing over 10 years [IRR= 3.15, 95% CI 3.13–3.17]); five times the rate of hospitalisation [IRR= 5.48, 95% CI 5.45–5.51]); and between a six to twelve-fold higher odds of mortality [OR at the year 5th = 5.93, 95% CI 5.2–6.76 and OR at year 10th= 12.03, 95% IC 11.03–13.13] compared to those in the LRC (Table 3). Individuals in HRC lost an average of 10.6 years of life after becoming multimorbid and before reaching the UK’s life expectancy of 81 years old compared to an average of 0.4 years of life lost in LRC (Table 2).

In South Asians, the HRC had the same 3 leading conditions (painful conditions, hypertension and depression) as in White (Figure 2C), with other common conditions including dermatitis, enthesopathy and synovial disorders and constipation. However, in contrast to the White population, both the HRC and LRCs in South Asians were predominantly comprised of women (66% and 88%, respectively) and were at greater deprivation (29% and 27%, respectively) (Table 2). Compared to the LRC, the South Asians in the HRC had twice the rate of primary care consultation [IRR= 1.96, 95% CI 1.89–2.03], twice the rate of long-term prescribing [IRR= 2.57, 95% CI 2.51–2.64]; and three times the hospitalisation rates [IRR= 3.13, 95% CI 3.06–3.19]. Individuals in the HRC had 8 and 11 times the odds of mortality by the end of the years 5th and 10th [OR= 8.48, 95% CI 3.06– 23.51; OR= 10.86, 95% CI 5.71–20.66, respectively] (Table 3). They also lost an average of 18.5 years of life after becoming multimorbid and before reaching 81 years old compared to 0.8 years of life lost compared to South Asians in the LRC, an order of magnitude greater than the same comparison in Whites (Table 2).

In Blacks, in addition to the 3 leading conditions common to other ethnicities (hypertension, painful conditions and depression) we observed constipation and obesity as highly prevalent LTCs in the HRC (Figure 2D). The HRC comprised a greater number of LTCs and a similar proportion of deprived population, compared to the LRC, consistent with the finding in South Asians. Individuals in the HRC had higher rates of primary care consultation [IRR= 1.74, 95 % CI 1.67–1.81] and long-term prescribing [IRR= 2.34, 95% CI 2.27–2.41], but the magnitude of difference was less than that seen in the other ethnic groups. However, hospitalisation rates in the HRC vs LRC group were significantly greater [IRR= 5.47, 95 % CI 5.35–5.6] and the HRC group also had the highest odds of mortality by the end of the year 5th [OR= 17.15, 95 % CI 5.39–54.55] and 10th [OR= 8.32, 95 % CI 5.24–13.2]. Multimorbid Blacks in the HRC lost and average of 18.0 years of life compared to 1.3 years in the LRC, consistent with the findings from the South Asian population.

We observed different associations between the socioeconomic deprivation of our population with outcomes that were further varied by ethnicity. While living in socioeconomically deprived areas was associated with lower rates of primary care consultations for South Asians [IRR= 0.92, 95% CI 0.89–0.94] compared to their peers from more affluent areas, it was associated with higher rates for Whites [IRR= 1.03, 95% CI 1.02– 1.04] living in deprived areas compared to their wealthier peers. However, Whites, South Asians and Blacks living in socioeconomically deprived areas had similarly higher rates of hospitalisations [IRR= 1.14, 95% CI 1.13–1.14, IRR=1.19, 95% CI 1.17–1.21, and IRR=1.38, 95%CI 1.35–1.42, respectively] and long-term prescribing [IRR= 1.25, 95% CI 1.25–1.26, IRR= 1.20, 95% CI 1.18–1.23, and IRR= 1.16, 95% CI 1.12–1.20, respectively] compared to those in less deprived areas (Table 3).

## Discussion

To our knowledge, this is the first study to investigate early onset of multimorbidity and its variation by ethnicity in a large UK population based sample. By applying a data-driven approach across multiple long-term conditions, and combining this with measures of healthcare utilisation, mortality and years of life lost, we have built new understanding of the burden and impact of early onset multimorbidity at population scale. Our findings provide important justification to improve the prevention, recognition and management of multimorbidity in young and diverse populations.

We show that in a large, multimorbid UK population, approximately 40% developed multimorbidity early (aged 16-39 years). Black and South Asian populations were more likely to become multimorbid early than Whites. We built on these findings to demonstrate the impact of early onset multimorbidity using measures of healthcare utilisation (primary care consultations, hospitalisation), long-term prescription use, mortality and years of life lost. In doing so, we have demonstrated for the first time that South Asian and Black groups with early onset multimorbidity died younger and lost more years of life once they become multimorbid compared to the White group. Although it is well established that multimorbidity increases with age [4,7,37], we show that multimorbidity is highly prevalent in younger populations and disproportionately affects minority ethnic and socially deprived groups, highlighting the need for early interventions to prevent and manage multimorbidity in those populations.

Our observation that the Black population with early onset multimorbidity has the lowest rate of primary care consultations and long-term prescriptions but higher rates of hospitalisation and mortality compared to south Asian and White groups suggests that a lack of routine care could underlie these worse outcomes. However, similar to the Black population, South Asian groups had higher mortality and years of life lost than the White group, yet had the highest rate of primary care consultations across all groups. These findings highlight the importance of understanding the complex relationship between ethnicity, access to and uptake of healthcare in order to improve outcomes from multimorbidity. Previous reports [38–43] have suggested that structural racism may play a role in explaining poorer health outcomes for certain groups within the UK – highlighting less positive experiences of care, insufficient support from local services, poorer treatment outcomes, and a lack of confidence in self-management of multimorbidity among minority ethnic groups compared to White groups [38–43] which is likely to contribute to inequalities in the effective identification and management of multimorbidity, resulting in higher mortality and years of life lost.

We highlighted important differences in outcomes within ethnic groups associated with deprivation. South Asians living in areas of high socioeconomic deprivation had lower rates of primary care consultations, but higher rates of hospitalisation and long-term prescribing than their peers living in more affluent areas. Black groups living in areas of high socioeconomic deprivation had higher rates of hospitalisation and long-term prescribing than their peers living in more affluent areas. In contrast, White groups with high socioeconomic deprivation had higher rates of consultation, hospitalisation and long-term prescribing than their peers from affluent areas. Previous studies show that people with multimorbidity living in more deprived areas may receive poorer quality healthcare represented by shorter consultation times, poorer patient-centeredness and lower perceived GP empathy compared to those living in more affluent areas [38,44]. These findings demonstrate the intersecting influences of ethnicity and socioeconomic deprivation that require action from clinical and public health systems to tackle upstream determinants of health that contribute to these stark inequalities and poor outcomes from multimorbidity.

Mortality for Black individuals in the highest risk multimorbidity cluster was greater by the end of year 5 than by the end of year 10. This may suggest a survivor effect in which individuals with more severe health conditions do not reach older ages. Differences in socioeconomic deprivation did not explain the mortality differential between clusters of South Asian and Black individuals. This may be due to the similar distribution of socioeconomic deprivation across all clusters in those ethnic groups, which was different from the White group, where those in the highest risk cluster had greater levels of socioeconomic deprivation than those in the lowest risk cluster.

Our comprehensive inclusion of 204 long-term conditions and the application of data driven approaches allowed us to generate unique insights into the early onset of multimorbidity, and the contribution of conditions with varied prevalence, e.g. by ethnicity, that are excluded from most other multimorbidity studies (e.g. sickle cell disease, chronic viral hepatitis, polycystic ovarian syndrome, thalassemia). By linking clusters of LTCs to clinically important outcomes, we were able to identify ‘high risk’ clusters of multimorbidity, defined by higher frequency of primary and secondary care consultations, long-term prescribing, greater years of life lost and greater mortality. The highest risk clusters in each ethnic group showed some similarities: they largely comprised older people living in areas of high socioeconomic deprivation. Common to all high risk clusters was a range of high prevalence physical and mental health conditions, including hypertension, depression, painful conditions, type 2 diabetes and anxiety. However, ethnic differences between the high risk clusters were observed. The high risk cluster in Whites comprised predominantly men and included conditions not observed in the high risk clusters in South Asians and Blacks: COPD, CHD, CKD, hearing loss and venous or lymphatic disease. In contrast, the high risk clusters in Blacks and South Asians were predominantly comprised of women and included conditions not seen in Whites: allergic rhinitis, iron deficiency anaemia, menorrhagia and polymenorrhoea, migraine and PTSD.

Differences in health service use and long-term prescribing associated with multimorbidity may be related to the combination of conditions that lead one to seek health care as well as having potentially manageable and resolvable conditions. In addition, individuals with a given LTC are more likely to seek healthcare services which may result in multiple LTCs being detected over time [12]. Furthermore, the number of LTCs identified may be related to the duration of an individual’s data linkage (and therefore follow-up time) length of the individuals’ follow-ups and age, as individuals with longer follow-up time and older ages and older have had time and opportunity to have their health-related conditions detected.

Guidelines in the UK that address and manage multimorbidity do not yet include guidance for managing the accumulation of LTCs in individuals with an early onset, nor bring guidance on targeted health care for minoritized ethnic groups or socioeconomically deprived groups at high premature risk for poorer health outcomes such as hospitalisation and premature mortality [45]. Public health policies that aim to reduce multimorbidity should be applied in younger populations, and although universal, should increase targeting towards minority groups and the more socioeconomically disadvantaged population.

## Strengths and limitations

The major strength of our study is the large scale of the population studied and the application of data-driven analyses across a robustly-defined set of 204 LTCs, building significantly on the existing evidence base. The strength of this approach has enabled us to ensure we represent diseases that may be ethnically patterned and which may contribute differently to the significant burden of early onset multimorbidity. Our approach elucidates early onset clusters of multimorbidity that confer particularly high risk of poor outcomes, and the identification of these provides a rationale for developing improved clinical pathways for the prevention and management of multimorbidity.

Our analysis focused on patients rather than diseases as the unit of analysis allowing for a deeper understanding of patient groups that share patterns of conditions and may provide essential information for the development of clinical guidelines and pathways of care.

Although there is no ‘gold standard’ on the selection of multiple long-term conditions for multimorbidity studies, we have shown that multimorbidity definitions can be operationalised in electronic health records, and our efforts have contributed to enhancing robust reproducible methodology.

There are some limitations of our work. Residual confounding may be present as there are likely to be other unmeasured factors underlying the association between multimorbidity and poorer outcomes that we were unable to study in our analysis (e.g. educational level, and aetiological factors). Another limitation is that 13.4% of the multimorbid population had no ethnicity recorded and may be inherently different (likely to have non-random missing data) and have poorer health than those with an ethnic group recorded [46].

There are some limitations intrinsic to the use of electronic health records. The health conditions selected might be subject to misclassification due to unrecorded, miscoded, and undiagnosed diseases. The age of onset might not reflect the actual age at which a given condition was diagnosed, but the date when it was entered in the patient’s medical record. Additionally, the number of LTCs identified is related to the use of the health care service, as well as the length of follow-up time measured as the duration of data availability within their EHR. Individuals with longer follow-up time have had time and opportunity to have multimorbidity detected. Last but not least, there may be a time-related bias in the LTCs coding given that we included any diagnosis code ever recorded and therefore, there are fewer codes being recorded particularly prior to late 1990 when 96% of the general practices were using computerised record systems [47].

## Conclusion

Early onset multimorbidity is the most common form of multimorbidity among minority ethnic in the UK. By characterising rates of primary care consultation and hospitalisation, long-term prescribing, mortality and loss of life expectancy, we were able to identify clusters of multimorbidity that were particularly high risk. Across ethnicities, these high risk clusters were common in more socioeconomically deprived individuals and contained several common long-term physical and mental health conditions including hypertension and depression. However, the high risk clusters also demonstrated variability between ethnic groups by sex and conditions. We hypothesise that the worse outcomes from early onset multimorbidity in Black and socioeconomically deprived groups may, in part, be due to receiving poorer routine healthcare These findings emphasise the need to identify, prevent and manage multimorbidity early in the life course. Most health systems remain focused on single disease management, and our findings add further weight to calls to restructure healthcare provision to do so. Our work provides additional insights into the need to ensure these healthcare improvements are equitable and reach those from socioeconomically deprived and diverse groups who are disproportionately and more severely affected by multimorbidity.

## Data Availability

The study uses data from the Clinical Practice Research Datalink (CPRD). CPRD does not allow the sharing of patient-level data. The data specification for the CPRD data set is available at: https://www.cprd.com/sites/default/files/CPRD_GOLD_Full_Data_Specification_v2.0_0.pdf. Additional information on the IMD patient-level linkage data is available at: https://cprd.com/sites/default/files/Documentation_SmallAreaData_Patient_set18_v2.7.pdf. The code list used in this study are available at https://github.com/f-eto/MULTIPLY-Initiative

