## Supplementary File for "Ethnic differences in early onset multimorbidity and associations with health service use, long-term prescribing, years of life lost, and mortality: an observational study using person-level clustering in the UK Clinical Practice Research Datalink"

**Supporting information**

1. **Prevalence of the long-term conditions in the population with early onset of multimorbidity according to ethnic groups and clusters**

| **Table S1**. Prevalence of the 204 long-term conditions per 100 according to ethnic groups (White n = 777,906; South Asian n = 33,915; and Black or Black British n = 26,048). Population with early onset of multimorbidity (16 to 39 years old). | | | | | | |
| --- | --- | --- | --- | --- | --- | --- |
| **Long-term condition** | **White** | | **South Asian** | | **Black or Black British** | |
|  | **Count** | **Prevalence** | **Count** | **Prevalence** | **Count** | **Prevalence** |
| ADHD and hyperkinetic disorders | 4192 | 0.5 | 37 | 0.1 | 50 | 0.2 |
| Adrenal insufficiency and Addison's disease | 1631 | 0.2 | 57 | 0.2 | 47 | 0.2 |
| Alcohol dependence and related disease | 62518 | 8.0 | 1073 | 3.2 | 1020 | 3.9 |
| Allergic and chronic rhinitis | 166989 | 21.5 | 9384 | 27.7 | 7506 | 28.8 |
| Alopecia areata and scarring alopecia | 4962 | 0.6 | 555 | 1.6 | 233 | 0.9 |
| Ankylosing spondylitis | 3492 | 0.4 | 92 | 0.3 | 22 | 0.1 |
| Anxiety and phobia | 265640 | 34.1 | 5725 | 16.9 | 4194 | 16.1 |
| Aortic aneurysm | 2654 | 0.3 | 40 | 0.1 | 24 | 0.1 |
| Aplastic anaemias | 1742 | 0.2 | 78 | 0.2 | 84 | 0.3 |
| Asbestosis | 570 | 0.1 | 2 | 0.0 | 1 | 0.0 |
| Asthma | 194209 | 25.0 | 6718 | 19.8 | 4794 | 18.4 |
| Atrial fibrillation and flutter | 23830 | 3.1 | 329 | 1.0 | 314 | 1.2 |
| Autism and Asperger's syndrome | 3599 | 0.5 | 55 | 0.2 | 96 | 0.4 |
| Autoimmune liver disease | 985 | 0.1 | 50 | 0.1 | 29 | 0.1 |
| Barrett's oesophagus | 6409 | 0.8 | 88 | 0.3 | 30 | 0.1 |
| Bipolar affective disorder and mania | 15626 | 2.0 | 409 | 1.2 | 547 | 2.1 |
| Blistering autoimmune skin conditions | 867 | 0.1 | 40 | 0.1 | 22 | 0.1 |
| Bronchiectasis | 6093 | 0.8 | 190 | 0.6 | 116 | 0.4 |
| Cardiac conduction defects | 11377 | 1.5 | 303 | 0.9 | 205 | 0.8 |
| Cardiomyopathy other | 4038 | 0.5 | 121 | 0.4 | 184 | 0.7 |
| Cerebral palsy | 1623 | 0.2 | 33 | 0.1 | 42 | 0.2 |
| Cerebrovascular disease | 19273 | 2.5 | 488 | 1.4 | 401 | 1.5 |
| Cervical carcinoma in situ | 35038 | 4.5 | 358 | 1.1 | 957 | 3.7 |
| Cholelithiasis | 57340 | 7.4 | 1757 | 5.2 | 1231 | 4.7 |
| Chronic fatigue | 15450 | 2.0 | 229 | 0.7 | 135 | 0.5 |
| Chronic kidney disease | 28909 | 3.7 | 992 | 2.9 | 1119 | 4.3 |
| Chronic obstructive pulmonary disease | 32685 | 4.2 | 406 | 1.2 | 287 | 1.1 |
| Chronic sinusitis | 96459 | 12.4 | 2877 | 8.5 | 1599 | 6.1 |
| Chronic ulcer of the skin | 37983 | 4.9 | 921 | 2.7 | 630 | 2.4 |
| Chronic viral hepatitis | 8042 | 1.0 | 402 | 1.2 | 738 | 2.8 |
| Coeliac disease | 6331 | 0.8 | 290 | 0.9 | 37 | 0.1 |
| Collapsed vertebra | 2624 | 0.3 | 44 | 0.1 | 26 | 0.1 |
| Congenital cardiac disease | 7905 | 1.0 | 256 | 0.8 | 165 | 0.6 |
| Constipation | 140503 | 18.1 | 7834 | 23.1 | 5392 | 20.7 |
| Coronary heart disease | 45160 | 5.8 | 1808 | 5.3 | 714 | 2.7 |
| Crohn's disease | 9931 | 1.3 | 360 | 1.1 | 127 | 0.5 |
| Cystic fibrosis | 615 | 0.1 | 14 | 0.0 | 13 | 0.0 |
| Cystic renal disease | 1471 | 0.2 | 43 | 0.1 | 45 | 0.2 |
| Dementia | 7002 | 0.9 | 108 | 0.3 | 79 | 0.3 |
| Depression | 379238 | 48.8 | 8729 | 25.7 | 7466 | 28.7 |
| Dermatitis (atopic/contact/other/unspecified) | 232811 | 29.9 | 11477 | 33.8 | 6563 | 25.2 |
| Diabetic eye disease | 16503 | 2.1 | 1116 | 3.3 | 601 | 2.3 |
| Diabetic neurological complications | 4942 | 0.6 | 181 | 0.5 | 110 | 0.4 |
| Disorders of autonomic nervous system | 5642 | 0.7 | 157 | 0.5 | 87 | 0.3 |
| Diverticular disease of intestine | 44018 | 5.7 | 515 | 1.5 | 500 | 1.9 |
| Down's syndrome | 710 | 0.1 | 27 | 0.1 | 15 | 0.1 |
| Dysmenorrhoea | 61487 | 7.9 | 2642 | 7.8 | 2554 | 9.8 |
| Eating disorders | 19606 | 2.5 | 429 | 1.3 | 338 | 1.3 |
| End stage renal disease | 3828 | 0.5 | 232 | 0.7 | 258 | 1.0 |
| Endometriosis | 35014 | 4.5 | 1168 | 3.4 | 1048 | 4.0 |
| Enteropathic arthropathy | 163 | 0.0 | 3 | 0.0 | 2 | 0.0 |
| Enthesopathies and synovial disorders | 198472 | 25.5 | 6936 | 20.5 | 4288 | 16.5 |
| Epilepsy | 30703 | 3.9 | 710 | 2.1 | 640 | 2.5 |
| Erectile dysfunction | 33454 | 4.3 | 1602 | 4.7 | 939 | 3.6 |
| Female genital prolapse | 30768 | 4.0 | 818 | 2.4 | 526 | 2.0 |
| Female infertility | 60400 | 7.8 | 4516 | 13.3 | 3061 | 11.8 |
| Fibromyalgia | 18120 | 2.3 | 618 | 1.8 | 265 | 1.0 |
| Folate deficiency, with and without anaemia | 7073 | 0.9 | 574 | 1.7 | 299 | 1.1 |
| Fracture of hip | 13543 | 1.7 | 158 | 0.5 | 138 | 0.5 |
| Gastritis and duodenitis | 95997 | 12.3 | 4297 | 12.7 | 2714 | 10.4 |
| Gastrointestinal angiodysplasia | 857 | 0.1 | 21 | 0.1 | 10 | 0.0 |
| Gastro-oesophageal reflux disease | 132758 | 17.1 | 5943 | 17.5 | 3428 | 13.2 |
| Giant cell arteritis | 1080 | 0.1 | 43 | 0.1 | 10 | 0.0 |
| Glaucoma | 9071 | 1.2 | 346 | 1.0 | 403 | 1.5 |
| Glomerulonephritis and other nephritides | 8151 | 1.0 | 397 | 1.2 | 362 | 1.4 |
| Gout | 20843 | 2.7 | 616 | 1.8 | 348 | 1.3 |
| Hearing loss | 83457 | 10.7 | 2330 | 6.9 | 1132 | 4.3 |
| Heart failure | 17366 | 2.2 | 462 | 1.4 | 421 | 1.6 |
| Heart valve disease non-rheumatic | 16991 | 2.2 | 486 | 1.4 | 465 | 1.8 |
| Hidradenitis suppurativa | 6717 | 0.9 | 256 | 0.8 | 323 | 1.2 |
| HIV | 1254 | 0.2 | 28 | 0.1 | 714 | 2.7 |
| Hodgkin lymphoma | 1423 | 0.2 | 42 | 0.1 | 29 | 0.1 |
| Hyperparathyroidism | 2762 | 0.4 | 180 | 0.5 | 158 | 0.6 |
| Hyperplasia of prostate | 12666 | 1.6 | 315 | 0.9 | 183 | 0.7 |
| Hyperprolactinaemia and prolactinoma | 2880 | 0.4 | 199 | 0.6 | 238 | 0.9 |
| Hypertension | 141866 | 18.2 | 5344 | 15.8 | 5498 | 21.1 |
| Hypertrophic cardiomyopathy | 717 | 0.1 | 37 | 0.1 | 51 | 0.2 |
| Hypertrophy of nasal turbinates | 23189 | 3.0 | 1370 | 4.0 | 380 | 1.5 |
| Hypopituitarism | 1830 | 0.2 | 95 | 0.3 | 66 | 0.3 |
| Hyposplenism | 2423 | 0.3 | 51 | 0.2 | 71 | 0.3 |
| Immunodeficiencies | 958 | 0.1 | 27 | 0.1 | 20 | 0.1 |
| Infection of bones and joints | 7040 | 0.9 | 220 | 0.6 | 232 | 0.9 |
| Intervertebral disc disorders | 53495 | 6.9 | 1466 | 4.3 | 719 | 2.8 |
| Intracerebral haemorrhage | 2560 | 0.3 | 92 | 0.3 | 88 | 0.3 |
| Intracranial hypertension | 1823 | 0.2 | 36 | 0.1 | 100 | 0.4 |
| Iron deficiency with and without anaemia | 57563 | 7.4 | 6763 | 19.9 | 4007 | 15.4 |
| Irritable bowel syndrome | 118879 | 15.3 | 3171 | 9.3 | 1887 | 7.2 |
| Juvenile arthritis | 828 | 0.1 | 20 | 0.1 | 17 | 0.1 |
| Learning disability | 10705 | 1.4 | 250 | 0.7 | 328 | 1.3 |
| Leukaemia | 2059 | 0.3 | 68 | 0.2 | 41 | 0.2 |
| Lichen planus | 5526 | 0.7 | 578 | 1.7 | 199 | 0.8 |
| Liver failure and transplant | 3921 | 0.5 | 98 | 0.3 | 84 | 0.3 |
| Liver fibrosis, sclerosis and cirrhosis | 6573 | 0.8 | 183 | 0.5 | 138 | 0.5 |
| Lupus erythematosus (local and systemic) | 2952 | 0.4 | 219 | 0.6 | 263 | 1.0 |
| Macular degeneration | 4764 | 0.6 | 114 | 0.3 | 86 | 0.3 |
| Male infertility | 52944 | 6.8 | 4030 | 11.9 | 2632 | 10.1 |
| Meniere disease | 3415 | 0.4 | 66 | 0.2 | 34 | 0.1 |
| Menorrhagia and polymenorrhoea | 135705 | 17.4 | 5877 | 17.3 | 5470 | 21.0 |
| Migraine | 128206 | 16.5 | 4764 | 14.0 | 3313 | 12.7 |
| Motor neuron disease | 482 | 0.1 | 16 | 0.0 | 5 | 0.0 |
| Multiple myeloma and malignant plasma cell neoplasms | 694 | 0.1 | 20 | 0.1 | 36 | 0.1 |
| Multiple sclerosis | 5317 | 0.7 | 119 | 0.4 | 97 | 0.4 |
| Myasthenia gravis | 458 | 0.1 | 15 | 0.0 | 29 | 0.1 |
| Myelodysplastic syndromes | 587 | 0.1 | 25 | 0.1 | 29 | 0.1 |
| Nasal polyp | 13682 | 1.8 | 704 | 2.1 | 333 | 1.3 |
| Neuromuscular dysfunction of bladder | 11544 | 1.5 | 427 | 1.3 | 300 | 1.2 |
| Non-acute cystitis | 4387 | 0.6 | 118 | 0.3 | 76 | 0.3 |
| Non-alcoholic fatty liver disease and steatohepatitis | 18774 | 2.4 | 1105 | 3.3 | 420 | 1.6 |
| Non-diabetic peripheral neuropathies (excluding cranial nerves and carpal tunnel syndrome) | 28106 | 3.6 | 783 | 2.3 | 542 | 2.1 |
| Non-Hodgkin lymphoma | 2830 | 0.4 | 82 | 0.2 | 71 | 0.3 |
| Non-malignant tumour of brain, central nervous system and pituitary | 3816 | 0.5 | 156 | 0.5 | 164 | 0.6 |
| Obesity | 137033 | 17.6 | 5435 | 16.0 | 5783 | 22.2 |
| Obsessive-compulsive disorder | 13229 | 1.7 | 254 | 0.7 | 115 | 0.4 |
| Obstructive and reflux uropathy | 12634 | 1.6 | 576 | 1.7 | 304 | 1.2 |
| Oesophagitis and oesophageal ulcer | 75035 | 9.6 | 2710 | 8.0 | 1381 | 5.3 |
| Osteoarthritis | 103125 | 13.3 | 2585 | 7.6 | 1871 | 7.2 |
| Osteoporosis | 20988 | 2.7 | 551 | 1.6 | 231 | 0.9 |
| Other anaemias | 74490 | 9.6 | 7517 | 22.2 | 5441 | 20.9 |
| Other haemolytic anaemias | 1421 | 0.2 | 229 | 0.7 | 295 | 1.1 |
| Other interstitial pulmonary diseases with fibrosis | 2433 | 0.3 | 102 | 0.3 | 81 | 0.3 |
| Other psychoactive substance misuse | 54107 | 7.0 | 797 | 2.3 | 1125 | 4.3 |
| Painful conditions | 171282 | 22.0 | 7057 | 20.8 | 4538 | 17.4 |
| Pancreatitis | 11291 | 1.5 | 384 | 1.1 | 256 | 1.0 |
| Parkinson's disease | 1966 | 0.3 | 42 | 0.1 | 16 | 0.1 |
| Peptic ulcer disease | 32208 | 4.1 | 1051 | 3.1 | 749 | 2.9 |
| Peripheral arterial disease | 12016 | 1.5 | 247 | 0.7 | 167 | 0.6 |
| Peripheral venous and lymphatic disease | 68961 | 8.9 | 2123 | 6.3 | 1047 | 4.0 |
| Personality disorder | 23619 | 3.0 | 252 | 0.7 | 382 | 1.5 |
| Polycystic ovarian syndrome | 19069 | 2.5 | 1610 | 4.7 | 779 | 3.0 |
| Polycythaemia vera | 1188 | 0.2 | 34 | 0.1 | 15 | 0.1 |
| Polymyalgia rheumatica | 3943 | 0.5 | 84 | 0.2 | 34 | 0.1 |
| Portal hypertension and oesophageal varices | 4369 | 0.6 | 100 | 0.3 | 60 | 0.2 |
| Post-traumatic stress and stress-related disorders | 172831 | 22.2 | 5395 | 15.9 | 4810 | 18.5 |
| Primary malignancy biliary tract | 391 | 0.1 | 7 | 0.0 | 6 | 0.0 |
| Primary malignancy bladder | 3649 | 0.5 | 73 | 0.2 | 52 | 0.2 |
| Primary malignancy bone and articular cartilage | 939 | 0.1 | 36 | 0.1 | 30 | 0.1 |
| Primary malignancy brain, other CNS and intracranial | 1786 | 0.2 | 52 | 0.2 | 33 | 0.1 |
| Primary malignancy breast | 15231 | 2.0 | 328 | 1.0 | 331 | 1.3 |
| Primary malignancy cervical | 11509 | 1.5 | 69 | 0.2 | 169 | 0.6 |
| Primary malignancy colorectal and anus | 4883 | 0.6 | 82 | 0.2 | 69 | 0.3 |
| Primary malignancy kidney and ureter | 3667 | 0.5 | 67 | 0.2 | 57 | 0.2 |
| Primary malignancy liver | 519 | 0.1 | 12 | 0.0 | 16 | 0.1 |
| Primary malignancy lung and trachea | 3953 | 0.5 | 49 | 0.1 | 59 | 0.2 |
| Primary malignancy malignant melanoma | 5878 | 0.8 | 25 | 0.1 | 15 | 0.1 |
| Primary malignancy mesothelioma | 155 | 0.0 | 0 | 0.0 | 0 | 0.0 |
| Primary malignancy oesophageal | 1004 | 0.1 | 10 | 0.0 | 12 | 0.0 |
| Primary malignancy oro-pharyngeal | 1951 | 0.3 | 53 | 0.2 | 33 | 0.1 |
| Primary malignancy other skin and subcutaneous tissue | 16570 | 2.1 | 46 | 0.1 | 31 | 0.1 |
| Primary malignancy others | 23950 | 3.1 | 420 | 1.2 | 356 | 1.4 |
| Primary malignancy ovarian | 1728 | 0.2 | 55 | 0.2 | 30 | 0.1 |
| Primary malignancy pancreatic | 879 | 0.1 | 14 | 0.0 | 25 | 0.1 |
| Primary malignancy prostate | 3244 | 0.4 | 35 | 0.1 | 71 | 0.3 |
| Primary malignancy stomach | 1177 | 0.2 | 25 | 0.1 | 27 | 0.1 |
| Primary malignancy testicular | 1741 | 0.2 | 33 | 0.1 | 16 | 0.1 |
| Primary malignancy thyroid | 10795 | 1.4 | 198 | 0.6 | 124 | 0.5 |
| Primary malignancy uterine | 1426 | 0.2 | 61 | 0.2 | 30 | 0.1 |
| Psoriasis | 46855 | 6.0 | 1314 | 3.9 | 352 | 1.4 |
| Psoriatic arthropathy | 4858 | 0.6 | 145 | 0.4 | 21 | 0.1 |
| Ptosis of eyelid | 3896 | 0.5 | 136 | 0.4 | 121 | 0.5 |
| Pulmonary hypertension | 2349 | 0.3 | 84 | 0.2 | 165 | 0.6 |
| Respiratory failure | 10680 | 1.4 | 263 | 0.8 | 279 | 1.1 |
| Retinal detachments and breaks | 6028 | 0.8 | 196 | 0.6 | 189 | 0.7 |
| Retinal vascular occlusions | 1855 | 0.2 | 65 | 0.2 | 48 | 0.2 |
| Rheumatic valve disease | 319 | 0.0 | 27 | 0.1 | 28 | 0.1 |
| Rheumatoid arthritis | 15148 | 1.9 | 693 | 2.0 | 386 | 1.5 |
| Rosacea | 32097 | 4.1 | 934 | 2.8 | 334 | 1.3 |
| Sarcoidosis | 2670 | 0.3 | 162 | 0.5 | 206 | 0.8 |
| SARS-CoV-2 | 6606 | 0.8 | 520 | 1.5 | 252 | 1.0 |
| Schizophrenia and non-organic psychosis | 21319 | 2.7 | 853 | 2.5 | 1505 | 5.8 |
| Scoliosis | 11214 | 1.4 | 234 | 0.7 | 242 | 0.9 |
| Seborrheic dermatitis | 47623 | 6.1 | 2674 | 7.9 | 1186 | 4.6 |
| Secondary malignancy and metastasis | 10726 | 1.4 | 198 | 0.6 | 228 | 0.9 |
| Secondary polycythaemia | 1564 | 0.2 | 45 | 0.1 | 21 | 0.1 |
| Sick sinus syndrome | 829 | 0.1 | 15 | 0.0 | 20 | 0.1 |
| Sickle-cell anaemia | 162 | 0.0 | 56 | 0.2 | 718 | 2.8 |
| Sjogren's disease | 1601 | 0.2 | 110 | 0.3 | 66 | 0.3 |
| Sleep apnoea | 18558 | 2.4 | 691 | 2.0 | 640 | 2.5 |
| Somatoform and dissociative disorders | 50369 | 6.5 | 2398 | 7.1 | 1532 | 5.9 |
| Spina bifida | 2680 | 0.3 | 41 | 0.1 | 34 | 0.1 |
| Spinal stenosis | 9994 | 1.3 | 321 | 0.9 | 188 | 0.7 |
| Spondylolisthesis | 5313 | 0.7 | 103 | 0.3 | 69 | 0.3 |
| Spondylosis | 42135 | 5.4 | 1237 | 3.6 | 608 | 2.3 |
| Subarachnoid haemorrhage | 3957 | 0.5 | 91 | 0.3 | 90 | 0.3 |
| Subdural haematoma | 2869 | 0.4 | 68 | 0.2 | 71 | 0.3 |
| Supraventricular tachycardia | 10342 | 1.3 | 275 | 0.8 | 193 | 0.7 |
| Systemic sclerosis | 572 | 0.1 | 28 | 0.1 | 25 | 0.1 |
| Thalassaemia | 822 | 0.1 | 572 | 1.7 | 349 | 1.3 |
| Thrombocytopenia primary, secondary and other | 8749 | 1.1 | 371 | 1.1 | 494 | 1.9 |
| Thrombophilia | 6674 | 0.9 | 207 | 0.6 | 164 | 0.6 |
| Thyroid disease | 64220 | 8.3 | 3925 | 11.6 | 1438 | 5.5 |
| Tinnitus | 32166 | 4.1 | 1180 | 3.5 | 695 | 2.7 |
| Trigeminal neuralgia | 6675 | 0.9 | 172 | 0.5 | 111 | 0.4 |
| Tuberculosis | 4954 | 0.6 | 1593 | 4.7 | 916 | 3.5 |
| Type 1 diabetes | 5453 | 0.7 | 95 | 0.3 | 108 | 0.4 |
| Type 2 diabetes | 42951 | 5.5 | 3761 | 11.1 | 1896 | 7.3 |
| Ulcerative colitis | 11884 | 1.5 | 547 | 1.6 | 164 | 0.6 |
| Unspecified or rare diabetes | 9446 | 1.2 | 598 | 1.8 | 452 | 1.7 |
| Urinary incontinence | 55033 | 7.1 | 2115 | 6.2 | 1307 | 5.0 |
| Urolithiasis | 37184 | 4.8 | 1673 | 4.9 | 719 | 2.8 |
| Urticaria | 54029 | 6.9 | 3122 | 9.2 | 1550 | 6.0 |
| Venous thromboembolism | 30667 | 3.9 | 660 | 1.9 | 793 | 3.0 |
| Visual impairment and blindness | 16462 | 2.1 | 611 | 1.8 | 572 | 2.2 |
| Vitamin B12 deficiency with and without anaemia | 27777 | 3.6 | 2649 | 7.8 | 407 | 1.6 |
| Vitiligo | 3786 | 0.5 | 463 | 1.4 | 208 | 0.8 |
| ADHD: attention deficit hyperactivity disorder, CNS: central nervous system, HIV: human immunodeficiency virus, SARS-CoV-2: severe acute respiratory syndrome coronavirus 2. | | | | | | |

| **Table S2**. Prevalence of the 204 long-term conditions per 100 according to clusters within each ethnic group (White n = 777,906; South Asian n = 33,915; and Black or Black British n = 26,048). | | | | | | | | | | |
| --- | --- | --- | --- | --- | --- | --- | --- | --- | --- | --- |
| **Long-term condition** | **White** | | | | **South Asian** | | | **Black or Black British** | | |
|  | **Cluster 1** | **Cluster 2** | **Cluster 3** | **Cluster 4** | **Cluster 1** | **Cluster 2** | **Cluster 3** | **Cluster 1** | **Cluster 2** | **Cluster 3** |
| ADHD and hyperkinetic disorders | 0.7 | 0.3 | 0.3 | 0.2 | 0.1 | 0.1 | 0 | 0.2 | 0.3 | 0 |
| Adrenal insufficiency and Addison's disease | 0.1 | 0.3 | 0.9 | 0.1 | 0.1 | 0.5 | 0 | 0.1 | 0.7 | 0.1 |
| Alcohol dependence and related disease | 7.2 | 6.2 | 18.2 | 3 | 3.1 | 4.4 | 1.2 | 3.7 | 6.8 | 1.2 |
| Allergic and chronic rhinitis | 18.9 | 30.9 | 20.7 | 17.9 | 25.7 | 39.2 | 20.4 | 27.3 | 37.8 | 24.1 |
| Alopecia areata and scarring alopecia | 0.6 | 0.8 | 0.8 | 0.5 | 1.6 | 2.1 | 1.1 | 0.8 | 1.4 | 1 |
| Ankylosing spondylitis | 0.3 | 0.8 | 1.2 | 0.2 | 0.2 | 0.7 | 0.1 | 0.1 | 0.2 | 0 |
| Anxiety and phobia | 28 | 53.2 | 41.4 | 23.9 | 14 | 32.1 | 10.5 | 14.1 | 28.6 | 10.3 |
| Aortic aneurysm | 0 | 0.1 | 2.6 | 0 | 0 | 0.6 | 0 | 0 | 0.4 | 0 |
| Aplastic anaemias | 0.1 | 0.1 | 1.5 | 0.1 | 0.1 | 0.9 | 0.1 | 0.2 | 1.2 | 0.1 |
| Asbestosis | 0 | 0 | 0.6 | 0 | 0 | 0 | 0 | 0 | 0 | 0 |
| Asthma | 22.1 | 32.1 | 31.5 | 18.1 | 17.7 | 32.4 | 12 | 16.7 | 29.8 | 12.6 |
| Atrial fibrillation and flutter | 0.7 | 1.4 | 21.1 | 0.6 | 0.2 | 4.6 | 0.1 | 0.4 | 5.7 | 0.1 |
| Autism and Asperger's syndrome | 0.5 | 0.3 | 0.5 | 0.1 | 0.2 | 0.2 | 0 | 0.4 | 0.5 | 0 |
| Autoimmune liver disease | 0 | 0.1 | 0.7 | 0 | 0.1 | 0.6 | 0 | 0.1 | 0.4 | 0 |
| Barrett's oesophagus | 0.2 | 1.7 | 3.2 | 0.2 | 0.1 | 0.9 | 0 | 0 | 0.5 | 0 |
| Bipolar affective disorder and mania | 1.5 | 3 | 4.1 | 0.6 | 1 | 2.4 | 0.4 | 2 | 3.7 | 0.5 |
| Blistering autoimmune skin conditions | 0.1 | 0.1 | 0.4 | 0 | 0.1 | 0.3 | 0 | 0.1 | 0.2 | 0.1 |
| Bronchiectasis | 0.2 | 1 | 4.3 | 0.3 | 0.2 | 2 | 0.2 | 0.2 | 1.7 | 0.1 |
| Cardiac conduction defects | 0.5 | 0.8 | 8.9 | 0.4 | 0.3 | 3.8 | 0.2 | 0.4 | 3.4 | 0.1 |
| Cardiomyopathy other | 0.1 | 0.1 | 3.8 | 0.1 | 0.1 | 1.6 | 0.1 | 0.2 | 3.5 | 0.1 |
| Cerebral Palsy | 0.2 | 0.2 | 0.5 | 0 | 0.1 | 0.1 | 0 | 0.1 | 0.4 | 0 |
| Cerebrovascular disease | 0.6 | 2.3 | 14.7 | 0.7 | 0.4 | 6.5 | 0.2 | 0.5 | 7.3 | 0.4 |
| Cervical carcinoma in situ | 4.4 | 5.8 | 1.9 | 5.3 | 1 | 1.4 | 0.9 | 3.7 | 3.5 | 3.2 |
| Cholelithiasis | 4.1 | 14.8 | 14.2 | 5.7 | 3.6 | 12.4 | 3.9 | 3.5 | 11 | 4 |
| Chronic fatigue | 1.1 | 5.3 | 2 | 1.1 | 0.3 | 2.3 | 0.3 | 0.3 | 1.6 | 0.1 |
| Chronic Kidney Disease | 0.6 | 3.7 | 23.3 | 1.2 | 0.9 | 12.6 | 0.8 | 1.5 | 18.9 | 2.6 |
| Chronic obstructive pulmonary disease | 0.9 | 5.7 | 22.8 | 0.9 | 0.3 | 5.3 | 0.2 | 0.4 | 4.9 | 0.2 |
| Chronic sinusitis | 8.3 | 25.2 | 15.7 | 9.2 | 6.6 | 18.3 | 4.5 | 4.7 | 14 | 4.6 |
| Chronic ulcer of the skin | 2.5 | 5 | 20.1 | 2.2 | 1.7 | 8 | 0.7 | 1.3 | 9 | 0.8 |
| Chronic viral hepatitis | 0.8 | 0.6 | 3.1 | 0.5 | 1 | 2.2 | 0.9 | 2.7 | 3.6 | 2.1 |
| Coeliac disease | 0.6 | 1.4 | 1.1 | 0.5 | 0.8 | 1.3 | 0.6 | 0.1 | 0.2 | 0.1 |
| Collapsed vertebra | 0.1 | 0.3 | 2.1 | 0.1 | 0 | 0.6 | 0 | 0 | 0.4 | 0.1 |
| Congenital cardiac disease | 0.8 | 0.7 | 3.2 | 0.6 | 0.6 | 1.6 | 0.5 | 0.4 | 2 | 0.2 |
| Constipation | 10.5 | 33.6 | 37.3 | 13.5 | 18.4 | 43.2 | 19.6 | 17 | 39 | 18.4 |
| Coronary heart disease | 1.2 | 5.5 | 36 | 1.2 | 1.5 | 23.6 | 1.1 | 0.7 | 14.1 | 0.7 |
| Crohn's disease | 1 | 1.9 | 2 | 0.7 | 0.9 | 1.9 | 0.6 | 0.4 | 1.1 | 0.2 |
| Cystic Fibrosis | 0 | 0.1 | 0.2 | 0.1 | 0 | 0.1 | 0 | 0 | 0.2 | 0 |
| Cystic renal disease | 0.1 | 0.1 | 0.8 | 0.1 | 0.1 | 0.5 | 0 | 0.1 | 0.6 | 0.1 |
| Dementia | 0.1 | 0.4 | 7.1 | 0.1 | 0 | 1.7 | 0 | 0.1 | 1.6 | 0.1 |
| Depression | 40.9 | 72 | 58.8 | 35.8 | 21.1 | 48.9 | 17.1 | 25.5 | 49.2 | 19.1 |
| Dermatitis (atopic/contact/other/unspecified) | 26.1 | 40 | 34.1 | 27.3 | 30.8 | 48.2 | 28.4 | 23.3 | 35.4 | 21.3 |
| Diabetic eye disease | 0.7 | 1.2 | 12.9 | 0.6 | 1 | 14.2 | 0.8 | 0.6 | 11.2 | 1.2 |
| Diabetic neurological complications | 0 | 0.1 | 5.5 | 0 | 0 | 3 | 0 | 0 | 2.6 | 0.1 |
| Disorders of autonomic nervous system | 0.3 | 1.3 | 2.7 | 0.2 | 0.2 | 1.9 | 0.1 | 0.1 | 1.7 | 0.1 |
| Diverticular disease of intestine | 1.7 | 11.5 | 20.5 | 2.6 | 0.6 | 5.9 | 0.5 | 0.7 | 8.2 | 1.2 |
| Down's syndrome | 0.1 | 0.1 | 0.2 | 0.1 | 0 | 0.2 | 0.1 | 0 | 0.2 | 0 |
| Dysmenorrhoea | 5.8 | 17 | 2.8 | 10.8 | 6.2 | 12.4 | 9.9 | 8.1 | 14.2 | 14.9 |
| Eating disorders | 2 | 3.8 | 3.6 | 1.8 | 1.1 | 2.4 | 0.7 | 1.1 | 2.7 | 0.7 |
| End stage renal disease | 0.1 | 0 | 4 | 0 | 0.1 | 3.7 | 0 | 0.1 | 6.1 | 0.2 |
| Endometriosis | 2.7 | 9.7 | 1.6 | 11.2 | 2.1 | 5.7 | 7.8 | 2.7 | 6.4 | 9.7 |
| Enteropathic arthropathy | 0 | 0 | 0.1 | 0 | 0 | 0 | 0 | 0 | 0 | 0 |
| Enthesopathies and synovial disorders | 16.9 | 46.7 | 41.9 | 19.9 | 14.4 | 47.9 | 14.5 | 12 | 37.1 | 16.5 |
| Epilepsy | 3 | 3.9 | 10.4 | 1.6 | 1.7 | 4.2 | 1 | 1.9 | 6.2 | 0.8 |
| Erectile dysfunction | 2.8 | 3 | 17 | 1.2 | 3.2 | 12.9 | 1.6 | 2.8 | 8.7 | 1.4 |
| Female genital prolapse | 1.5 | 10.9 | 6.6 | 3.3 | 1.2 | 7.7 | 1.5 | 1.3 | 6.1 | 1.4 |
| Female infertility | 0 | 4.9 | 4.1 | 99.9 | 0 | 9.5 | 99.9 | 0 | 8.1 | 99.9 |
| Fibromyalgia | 0.5 | 8.8 | 2.8 | 0.8 | 0.5 | 7.5 | 1.1 | 0.3 | 4.8 | 0.2 |
| Folate deficiency, with and without anaemia | 0.4 | 1.7 | 2.9 | 0.5 | 1.3 | 3.4 | 1.4 | 0.8 | 3.1 | 0.7 |
| Fracture of hip | 1.3 | 0.8 | 6.1 | 0.7 | 0.4 | 1 | 0.3 | 0.5 | 0.9 | 0.3 |
| Gastritis and duodenitis | 6.5 | 23.9 | 29.8 | 6 | 8.4 | 33.7 | 6.3 | 7.7 | 25.9 | 6.7 |
| Gastrointestinal angiodysplasia | 0 | 0.2 | 0.6 | 0 | 0 | 0.3 | 0 | 0 | 0.2 | 0 |
| Gastro-oesophageal reflux disease | 9.2 | 36.5 | 33.3 | 9.7 | 12.2 | 42.2 | 11.5 | 10 | 30.2 | 10 |
| Giant Cell arteritis | 0 | 0.2 | 0.8 | 0 | 0 | 0.7 | 0 | 0 | 0.2 | 0 |
| Glaucoma | 0.4 | 1.5 | 5.6 | 0.6 | 0.4 | 3.7 | 0.5 | 0.8 | 5.6 | 1.1 |
| Glomerulonephritis and other nephritides | 0.4 | 0.8 | 5.7 | 0.3 | 0.4 | 4.7 | 0.2 | 0.4 | 6.8 | 0.2 |
| Gout | 1.4 | 2.3 | 11.6 | 0.8 | 1 | 5.9 | 0.4 | 0.6 | 5.3 | 0.6 |
| Hearing loss | 7.6 | 15.5 | 22.1 | 7.7 | 5 | 15.6 | 4.5 | 3.4 | 9.9 | 3 |
| Heart failure | 0.1 | 0.2 | 19.1 | 0.2 | 0.1 | 7.2 | 0.2 | 0.2 | 9.5 | 0.3 |
| Heart valve disease non-rheumatic | 0.6 | 1.1 | 14.3 | 0.7 | 0.4 | 6.3 | 0.4 | 0.6 | 8.5 | 0.5 |
| Hidradenitis suppurativa | 0.7 | 1.6 | 0.9 | 0.7 | 0.7 | 1.2 | 0.5 | 1.1 | 2.1 | 0.7 |
| HIV | 0.2 | 0.1 | 0.2 | 0.1 | 0.1 | 0.1 | 0 | 3 | 1.3 | 3 |
| Hodgkin Lymphoma | 0.2 | 0.1 | 0.6 | 0.1 | 0.1 | 0.3 | 0 | 0.1 | 0.2 | 0 |
| Hyperparathyroidism | 0.1 | 0.5 | 1.8 | 0.2 | 0.2 | 2.2 | 0.2 | 0.2 | 2.9 | 0.4 |
| Hyperplasia of prostate | 0.5 | 1.2 | 10 | 0.2 | 0.2 | 4.4 | 0.1 | 0.3 | 3.1 | 0.1 |
| Hyperprolactinaemia and prolactinoma | 0.2 | 0.6 | 0.4 | 0.8 | 0.4 | 0.8 | 1.2 | 0.6 | 1.8 | 1.9 |
| Hypertension | 7.3 | 28.2 | 70.3 | 9 | 7.5 | 54.6 | 7.1 | 13.3 | 59.7 | 18.6 |
| Hypertrophic Cardiomyopathy | 0 | 0 | 0.6 | 0 | 0.1 | 0.3 | 0 | 0.1 | 0.9 | 0 |
| Hypertrophy of nasal turbinates | 2.9 | 3.9 | 2.7 | 1.7 | 4.2 | 4.6 | 1.9 | 1.4 | 2 | 1 |
| Hypopituitarism | 0.1 | 0.3 | 0.8 | 0.2 | 0.2 | 0.7 | 0.4 | 0.1 | 0.9 | 0.2 |
| Hyposplenism | 0.2 | 0.2 | 1.5 | 0.1 | 0.1 | 0.4 | 0.1 | 0.1 | 1.1 | 0.1 |
| Immunodeficiencies | 0 | 0.1 | 0.5 | 0 | 0 | 0.2 | 0 | 0 | 0.3 | 0 |
| Infection of bones and joints | 0.4 | 0.7 | 4.4 | 0.2 | 0.4 | 1.9 | 0.3 | 0.5 | 3 | 0.3 |
| Intervertebral disc disorders | 3.3 | 15.7 | 14.5 | 3.7 | 2.5 | 13.2 | 1.6 | 1.4 | 10.1 | 1.6 |
| Intracerebral haemorrhage | 0.1 | 0.2 | 1.9 | 0.1 | 0.1 | 1.2 | 0 | 0.2 | 1.2 | 0.2 |
| Intracranial hypertension | 0.2 | 0.4 | 0.2 | 0.2 | 0.1 | 0.3 | 0.1 | 0.3 | 1.1 | 0.1 |
| Iron deficiency with and without anaemia | 4.1 | 12.7 | 17.9 | 5.9 | 15.8 | 37.7 | 17 | 12.6 | 27.5 | 16 |
| Irritable bowel syndrome | 10.6 | 32.3 | 14.1 | 12.8 | 7.1 | 19.2 | 7.4 | 5.9 | 14.4 | 5.6 |
| Juvenile arthritis | 0.1 | 0.2 | 0.2 | 0.1 | 0 | 0.1 | 0 | 0 | 0.2 | 0 |
| Learning disability | 1.2 | 1.1 | 3.1 | 0.3 | 0.7 | 1.4 | 0.2 | 1.2 | 2.4 | 0 |
| Leukaemia | 0.1 | 0.2 | 1.2 | 0.2 | 0.1 | 0.7 | 0 | 0.1 | 0.5 | 0 |
| Lichen planus | 0.4 | 1.3 | 1.6 | 0.5 | 1.2 | 4.1 | 1.3 | 0.6 | 1.8 | 0.5 |
| Liver failure and transplant | 0.1 | 0.1 | 3.8 | 0.1 | 0.1 | 1.2 | 0 | 0.1 | 1.5 | 0.1 |
| Liver fibrosis, sclerosis and cirrhosis | 0.2 | 0.3 | 6.3 | 0.1 | 0.2 | 2.3 | 0.1 | 0.2 | 2.3 | 0.1 |
| Lupus erythematosus (local and systemic) | 0.2 | 0.8 | 1 | 0.2 | 0.4 | 1.9 | 0.3 | 0.5 | 3.9 | 0.4 |
| Macular degeneration | 0.1 | 0.5 | 4.1 | 0.2 | 0.1 | 1.5 | 0 | 0.1 | 1.5 | 0.2 |
| Male infertility | 0.2 | 3.2 | 4 | 87.6 | 0.2 | 8.1 | 88.1 | 0.1 | 6 | 85.7 |
| Meniere disease | 0.2 | 1 | 1.1 | 0.2 | 0.1 | 0.7 | 0 | 0 | 0.5 | 0.1 |
| Menorrhagia and polymenorrhoea | 11.5 | 40 | 10.9 | 21.2 | 13.3 | 31.3 | 19.8 | 16.8 | 34.6 | 29 |
| Migraine | 12.9 | 30.1 | 14.5 | 13.9 | 11.7 | 24.2 | 11.9 | 11.6 | 19.3 | 10 |
| Motor neuron disease | 0 | 0.1 | 0.2 | 0 | 0 | 0.2 | 0 | 0 | 0.1 | 0 |
| Multiple myeloma and malignant plasma cell neoplasms | 0 | 0.1 | 0.6 | 0 | 0 | 0.2 | 0 | 0 | 0.7 | 0 |
| Multiple sclerosis | 0.4 | 1.3 | 1.2 | 0.5 | 0.3 | 0.5 | 0.3 | 0.2 | 1.3 | 0.1 |
| Myasthenia gravis | 0 | 0.1 | 0.2 | 0 | 0 | 0.2 | 0 | 0.1 | 0.3 | 0.1 |
| Myelodysplastic syndromes | 0 | 0 | 0.5 | 0 | 0 | 0.4 | 0 | 0.1 | 0.4 | 0.1 |
| Nasal polyp | 1.3 | 2.7 | 3.2 | 1 | 2 | 3.1 | 1 | 1.1 | 2.2 | 0.8 |
| Neuromuscular dysfunction of bladder | 0.5 | 3.7 | 3.7 | 0.7 | 0.6 | 4.6 | 0.4 | 0.5 | 4.2 | 0.9 |
| Non-acute cystitis | 0.2 | 1.2 | 1.4 | 0.4 | 0.2 | 1 | 0.3 | 0.1 | 1.1 | 0.3 |
| Non-alcoholic fatty liver disease and steatohepatitis | 0.9 | 4.3 | 8.6 | 1 | 1.9 | 9.7 | 1.5 | 0.8 | 5.8 | 1.2 |
| Non-diabetic peripheral neuropathies (excluding cranial nerves and carpal tunnel syndrome) | 1.5 | 6 | 12.9 | 1.7 | 1.1 | 8.1 | 0.8 | 1 | 8.1 | 1.1 |
| Non-Hodgkin Lymphoma | 0.2 | 0.3 | 1.6 | 0.2 | 0.1 | 0.7 | 0.1 | 0.2 | 0.7 | 0.1 |
| Non-malignant tumour of brain, central nervous system and pituitary | 0.3 | 0.8 | 1.2 | 0.4 | 0.3 | 1.2 | 0.3 | 0.4 | 1.8 | 0.7 |
| Obesity | 12 | 29.5 | 32.3 | 12.8 | 12.4 | 31.2 | 14.4 | 18.7 | 37.9 | 22.4 |
| Obsessive-compulsive disorder | 1.4 | 2.9 | 2 | 0.9 | 0.7 | 1.2 | 0.5 | 0.4 | 0.7 | 0.3 |
| Obstructive and reflux uropathy | 1.1 | 1.7 | 4.9 | 0.9 | 1.4 | 3.3 | 0.8 | 0.8 | 3.2 | 1 |
| Oesophagitis and oesophageal ulcer | 4.3 | 21.4 | 23.8 | 4 | 4.7 | 23.8 | 4.2 | 3.4 | 15.6 | 3.6 |
| Osteoarthritis | 4.2 | 29.9 | 41.4 | 6.5 | 2.5 | 31.4 | 2.6 | 3.2 | 28.2 | 4.3 |
| Osteoporosis | 0.4 | 4.4 | 13.9 | 1.2 | 0.4 | 7.3 | 0.7 | 0.2 | 4.2 | 0.7 |
| Other anaemias | 5.4 | 14.9 | 26 | 7.6 | 17.6 | 41.3 | 19.4 | 17.2 | 37.3 | 21.4 |
| Other haemolytic anaemias | 0.1 | 0.1 | 0.5 | 0.1 | 0.6 | 0.9 | 0.6 | 0.9 | 2.5 | 0.7 |
| Other interstitial pulmonary diseases with fibrosis | 0 | 0.2 | 2.3 | 0 | 0.1 | 1.4 | 0 | 0.1 | 1.7 | 0.1 |
| Other psychoactive substance misuse | 6.7 | 6.4 | 11.3 | 2.3 | 2.4 | 3.4 | 0.5 | 4.4 | 5.7 | 1.2 |
| Painful conditions | 9 | 50.1 | 56.8 | 10 | 11.9 | 62.3 | 11.6 | 10.8 | 51.5 | 13.6 |
| Pancreatitis | 0.8 | 1.7 | 5.5 | 0.6 | 0.7 | 3.1 | 0.4 | 0.7 | 3 | 0.3 |
| Parkinson's disease | 0 | 0.2 | 1.7 | 0.1 | 0 | 0.6 | 0 | 0 | 0.4 | 0 |
| Peptic ulcer disease | 1.4 | 7 | 17.1 | 1 | 1.4 | 11.3 | 0.8 | 1.9 | 8.3 | 1.5 |
| Peripheral arterial disease | 0.2 | 0.7 | 11.6 | 0.2 | 0.1 | 3.5 | 0.1 | 0.1 | 3.6 | 0.1 |
| Peripheral venous and lymphatic disease | 5.7 | 12.6 | 21.7 | 6.7 | 4.5 | 14.6 | 3.9 | 2.9 | 10.2 | 2.3 |
| Personality disorder | 2.2 | 4.7 | 6.6 | 0.7 | 0.6 | 1.6 | 0.3 | 1.3 | 3.3 | 0.2 |
| Polycystic ovarian syndrome | 2.1 | 2.7 | 0.6 | 7.8 | 3.8 | 2.8 | 13.1 | 2.5 | 2.3 | 7.3 |
| Polycythaemia vera | 0.1 | 0.1 | 0.9 | 0 | 0 | 0.3 | 0.1 | 0 | 0.2 | 0 |
| Polymyalgia Rheumatica | 0 | 1 | 2.7 | 0.1 | 0 | 1.3 | 0 | 0 | 0.7 | 0.1 |
| Portal hypertension and oesophageal varices | 0.1 | 0.1 | 4.4 | 0.1 | 0.1 | 1.4 | 0 | 0.1 | 1 | 0.1 |
| Post-traumatic stress and stress-related disorders | 16.9 | 40.9 | 21.4 | 19.5 | 13.5 | 27.1 | 12.4 | 15.8 | 30.7 | 17.8 |
| Primary malignancy biliary tract | 0 | 0 | 0.4 | 0 | 0 | 0.1 | 0 | 0 | 0.1 | 0 |
| Primary malignancy bladder | 0.1 | 0.2 | 3.2 | 0.2 | 0.1 | 1 | 0 | 0 | 1 | 0.1 |
| Primary malignancy bone and articular cartilage | 0.1 | 0.1 | 0.4 | 0.1 | 0.1 | 0.3 | 0.1 | 0.1 | 0.4 | 0 |
| Primary malignancy brain, other CNS and intracranial | 0.1 | 0.2 | 0.9 | 0.1 | 0.1 | 0.3 | 0 | 0.1 | 0.5 | 0 |
| Primary malignancy breast | 0.9 | 4.1 | 4.1 | 2.4 | 0.5 | 2.8 | 1.1 | 0.8 | 3.6 | 1.2 |
| Primary malignancy cervical | 1.4 | 1.8 | 1 | 1.4 | 0.2 | 0.4 | 0.2 | 0.7 | 0.7 | 0.3 |
| Primary malignancy colorectal and anus | 0.2 | 0.6 | 3.5 | 0.3 | 0.1 | 1.1 | 0 | 0.1 | 1.3 | 0 |
| Primary malignancy kidney and ureter | 0.1 | 0.2 | 3.2 | 0.2 | 0.1 | 0.9 | 0 | 0 | 1.2 | 0.1 |
| Primary malignancy liver | 0 | 0 | 0.5 | 0 | 0 | 0.2 | 0 | 0 | 0.2 | 0 |
| Primary malignancy lung and trachea | 0.1 | 0.4 | 3.5 | 0.1 | 0 | 0.6 | 0 | 0.1 | 1.1 | 0 |
| Primary malignancy malignant melanoma | 0.5 | 0.9 | 1.7 | 0.8 | 0 | 0.1 | 0.2 | 0 | 0.2 | 0.1 |
| Primary malignancy mesothelioma | 0 | 0 | 0.1 | 0 | NA | NA | NA | NA | NA | NA |
| Primary malignancy oesophageal | 0 | 0 | 1 | 0 | 0 | 0.1 | 0 | 0 | 0.2 | 0.1 |
| Primary malignancy oro-pharyngeal | 0.1 | 0.2 | 1.3 | 0.1 | 0.1 | 0.4 | 0 | 0.1 | 0.5 | 0 |
| Primary malignancy other skin and subcutaneous tissue | 1 | 2.7 | 8.1 | 1.8 | 0.1 | 0.4 | 0.1 | 0 | 0.5 | 0 |
| Primary malignancy others | 1 | 3.2 | 15.4 | 1.7 | 0.5 | 4.5 | 0.6 | 0.5 | 6.2 | 0.5 |
| Primary malignancy ovarian | 0.1 | 0.4 | 0.8 | 0.3 | 0.1 | 0.5 | 0.2 | 0 | 0.4 | 0.1 |
| Primary malignancy pancreatic | 0 | 0.1 | 0.8 | 0 | 0 | 0.2 | 0 | 0 | 0.6 | 0 |
| Primary malignancy prostate | 0.1 | 0.1 | 3 | 0 | 0 | 0.6 | 0 | 0.1 | 1.1 | 0.3 |
| Primary malignancy stomach | 0 | 0.1 | 1.2 | 0 | 0 | 0.4 | 0 | 0 | 0.4 | 0.2 |
| Primary malignancy testicular | 0.2 | 0 | 0.5 | 0.1 | 0.1 | 0.1 | 0 | 0 | 0.1 | 0 |
| Primary malignancy thyroid | 0.5 | 1.4 | 6.8 | 0.8 | 0.4 | 1.7 | 0.3 | 0.2 | 2.2 | 0.2 |
| Primary malignancy uterine | 0 | 0.4 | 0.7 | 0.2 | 0 | 0.8 | 0.2 | 0 | 0.6 | 0.1 |
| Psoriasis | 5 | 7.7 | 9.1 | 4.8 | 3.4 | 6.7 | 2.5 | 1.2 | 2.3 | 0.8 |
| Psoriatic arthropathy | 0.3 | 1.3 | 1.4 | 0.3 | 0.3 | 1.3 | 0.2 | 0 | 0.4 | 0 |
| Ptosis of eyelid | 0.3 | 0.8 | 1.4 | 0.3 | 0.3 | 1.1 | 0.1 | 0.3 | 1.5 | 0.1 |
| Pulmonary hypertension | 0 | 0 | 2.6 | 0 | 0 | 1.3 | 0 | 0.1 | 3.6 | 0.1 |
| Respiratory failure | 0.3 | 0.7 | 9.7 | 0.2 | 0.2 | 3.4 | 0.1 | 0.3 | 5.4 | 0.1 |
| Retinal detachments and breaks | 0.5 | 0.6 | 3.1 | 0.4 | 0.4 | 1.6 | 0.2 | 0.4 | 2.6 | 0.3 |
| Retinal vascular occlusions | 0.1 | 0.2 | 1.4 | 0.1 | 0.1 | 0.8 | 0.1 | 0 | 0.9 | 0 |
| Rheumatic valve disease | 0 | 0 | 0.3 | 0 | 0 | 0.4 | 0 | 0 | 0.6 | 0 |
| Rheumatoid Arthritis | 0.6 | 4.3 | 6.5 | 1 | 1 | 6.8 | 1.3 | 0.7 | 5.8 | 0.8 |
| Rosacea | 3.3 | 6.7 | 4.3 | 4.2 | 2.5 | 3.7 | 2.6 | 1.2 | 1.8 | 0.9 |
| Sarcoidosis | 0.2 | 0.5 | 1.1 | 0.2 | 0.3 | 1.4 | 0.1 | 0.5 | 2.4 | 0.5 |
| SARS-CoV-2 | 0.8 | 0.9 | 1.2 | 0.8 | 1.4 | 2.3 | 1.3 | 0.8 | 1.8 | 1.1 |
| Schizophrenia and non-organic psychosis | 2.4 | 2.5 | 6.5 | 0.5 | 2.3 | 4.5 | 0.8 | 5.8 | 8.6 | 1.2 |
| Scoliosis | 1 | 2.1 | 3.3 | 0.9 | 0.4 | 1.9 | 0.5 | 0.7 | 2.3 | 0.3 |
| Seborrheic dermatitis | 4.9 | 9.3 | 8 | 5.1 | 7.2 | 11.7 | 5.8 | 4.1 | 7.2 | 3.8 |
| Secondary malignancy and metastasis | 0.3 | 1.2 | 8.3 | 0.8 | 0.2 | 2.4 | 0.2 | 0.3 | 4.3 | 0.3 |
| Secondary polycythaemia | 0.1 | 0.1 | 1.1 | 0.1 | 0.1 | 0.5 | 0.1 | 0 | 0.3 | 0 |
| Sick sinus syndrome | 0 | 0.1 | 0.7 | 0 | 0 | 0.1 | 0 | 0 | 0.3 | 0 |
| Sickle-cell anaemia | 0 | 0 | 0 | 0 | 0.1 | 0.3 | 0.2 | 2.2 | 5.7 | 2.1 |
| Sjogren's disease | 0 | 0.6 | 0.6 | 0.1 | 0.1 | 1.4 | 0.1 | 0.1 | 1.3 | 0.1 |
| Sleep apnoea | 1.1 | 3.9 | 8.2 | 0.8 | 1.2 | 6.2 | 0.7 | 1.6 | 7.4 | 1.1 |
| Somatoform and dissociative disorders | 4 | 14.8 | 7.4 | 4.6 | 5.1 | 15.9 | 5.4 | 4.7 | 11.7 | 5.3 |
| Spina bifida | 0.2 | 0.6 | 0.7 | 0.2 | 0.1 | 0.3 | 0 | 0.1 | 0.4 | 0.1 |
| Spinal stenosis | 0.2 | 3.2 | 4.8 | 0.3 | 0.3 | 4.1 | 0.3 | 0.2 | 3.6 | 0.1 |
| Spondylolisthesis | 0.2 | 1.6 | 2.1 | 0.3 | 0.1 | 1.3 | 0.1 | 0.1 | 1 | 0.1 |
| Spondylosis | 1 | 14 | 18.5 | 1.9 | 1.2 | 15.1 | 1.1 | 0.7 | 11.1 | 0.9 |
| Subarachnoid haemorrhage | 0.3 | 0.4 | 1.9 | 0.2 | 0.2 | 0.8 | 0.1 | 0.2 | 0.9 | 0.2 |
| Subdural haematoma | 0.2 | 0.1 | 1.7 | 0.1 | 0.1 | 0.7 | 0 | 0.2 | 0.9 | 0.1 |
| Supraventricular tachycardia | 0.8 | 1.5 | 4.5 | 0.7 | 0.6 | 2.2 | 0.3 | 0.4 | 2.5 | 0.4 |
| Systemic sclerosis | 0 | 0.1 | 0.3 | 0.1 | 0 | 0.4 | 0 | 0 | 0.5 | 0 |
| Thalassaemia | 0.1 | 0.1 | 0.1 | 0.1 | 1.5 | 2.4 | 1.4 | 1.1 | 2.4 | 1.2 |
| Thrombocytopenia primary, secondary and other | 0.7 | 0.7 | 4.7 | 0.7 | 0.8 | 2.6 | 0.6 | 1.6 | 3.8 | 0.9 |
| Thrombophilia | 0.7 | 1.2 | 1 | 1.4 | 0.4 | 1 | 1 | 0.4 | 1.6 | 0.8 |
| Thyroid disease | 5 | 14.8 | 16 | 8 | 9.7 | 17.8 | 12.5 | 4.1 | 12.3 | 5.3 |
| Tinnitus | 2.6 | 7.5 | 7.5 | 3.1 | 2.4 | 8.5 | 2.4 | 1.8 | 6.4 | 2.9 |
| Trigeminal neuralgia | 0.3 | 2.3 | 1.6 | 0.5 | 0.3 | 1.6 | 0.3 | 0.2 | 1.4 | 0.2 |
| Tuberculosis | 0.2 | 0.6 | 3.3 | 0.2 | 3.6 | 9.6 | 3.5 | 3.5 | 3.8 | 3.1 |
| Type 1 Diabetes | 0.8 | 0.2 | 1.4 | 0.3 | 0.3 | 0.1 | 0.1 | 0.4 | 0.6 | 0.3 |
| Type 2 Diabetes | 1.6 | 8.3 | 25.5 | 2.1 | 5.6 | 36.3 | 6.3 | 3.9 | 24.7 | 5.6 |
| Ulcerative colitis | 1.3 | 1.9 | 2.5 | 1 | 1.5 | 2.7 | 0.7 | 0.6 | 1.3 | 0.2 |
| Unspecified or Rare Diabetes | 0.4 | 0.7 | 7.3 | 0.4 | 0.8 | 6.2 | 1 | 1 | 6 | 0.9 |
| Urinary Incontinence | 2.8 | 17.2 | 15.8 | 4.8 | 3.7 | 18.1 | 3.6 | 3 | 16 | 3.1 |
| Urolithiasis | 3.3 | 6.7 | 11 | 2.6 | 4 | 10.1 | 2.6 | 2.1 | 6.5 | 1.7 |
| Urticaria | 5.6 | 11.7 | 6.8 | 6.2 | 7.8 | 15.6 | 7.6 | 5.3 | 9.4 | 5.2 |
| Venous thromboembolism | 1.9 | 4.6 | 15.9 | 1.6 | 1.1 | 6.2 | 0.7 | 1.6 | 10.9 | 1.3 |
| Visual impairment and blindness | 1.2 | 2.6 | 7.3 | 0.9 | 1.2 | 4.9 | 0.9 | 1.4 | 7 | 0.6 |
| Vitamin B12 deficiency with and without anaemia | 1.9 | 6.7 | 8.5 | 2.3 | 5.7 | 17.4 | 5.9 | 1 | 4.5 | 1.4 |
| Vitiligo | 0.4 | 0.7 | 0.5 | 0.4 | 1.2 | 2.2 | 1 | 0.6 | 1.5 | 0.9 |
| ADHD: attention deficit hyperactivity disorder, CNS: central nervous system, HIV: human immunodeficiency virus, SARS-CoV-2: severe acute respiratory syndrome coronavirus 2. | | | | | | | | | | |

2. **Flow chart for the selection of our study population**

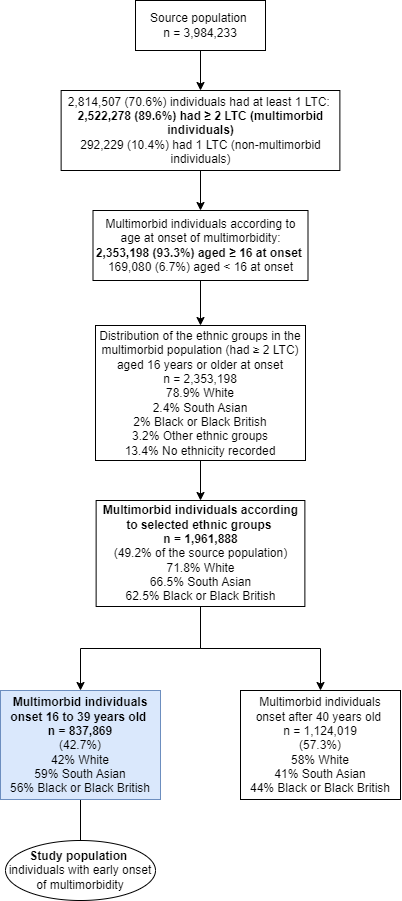

3. **Fit statistics for the model selection**

To select the models with the best class solution we verified the fit statistics (BIC, SABIC, likelihood ratio and entropy) for each model along with the clinical judgement of the most meaningful clusters among a couple of candidates for the best solution.

The BIC is considered the most reliable indicator of model fit (Nylund et al., 2007; Vermunt, 2002). Although, in practice, it is common that the BIC and SABIC (BIC adjusted for the sample size) decrease as you add classes to the model. Therefore, we opt for the most parsimonious model when evaluating these indexes. The likelihood ratio shows the difference in likelihood between the fitted model (e.g. *k* model being tested) and the *k-1* model, where *k* is the number of classes. Lower values indicate a more parsimonious model. The entropy assessed how accurately the classes classify individuals. In general, a value closer to 1 is desirable, although there is no agreed-upon cut-off criterion for entropy (B. O. Muthén, 2008). In practice, we inspected the plots for an “elbow” or a point where we do not see much improvement in the model fit (e.g. small decreases in the fit statistics for each additional class).

A 4-class model for the White population (Figure 1A, Table S2) and a 3-class model for both the South Asian and Black or Black British (Figure 1B and 1C, respectively, Table S2) were selected as the best solution given that only small improvements in the fit indexes for additional classes can be seen from upper classes. Additionally, the respective classes show an improvement in the model classification (entropy) compared to their neighbour models.

**A: White**

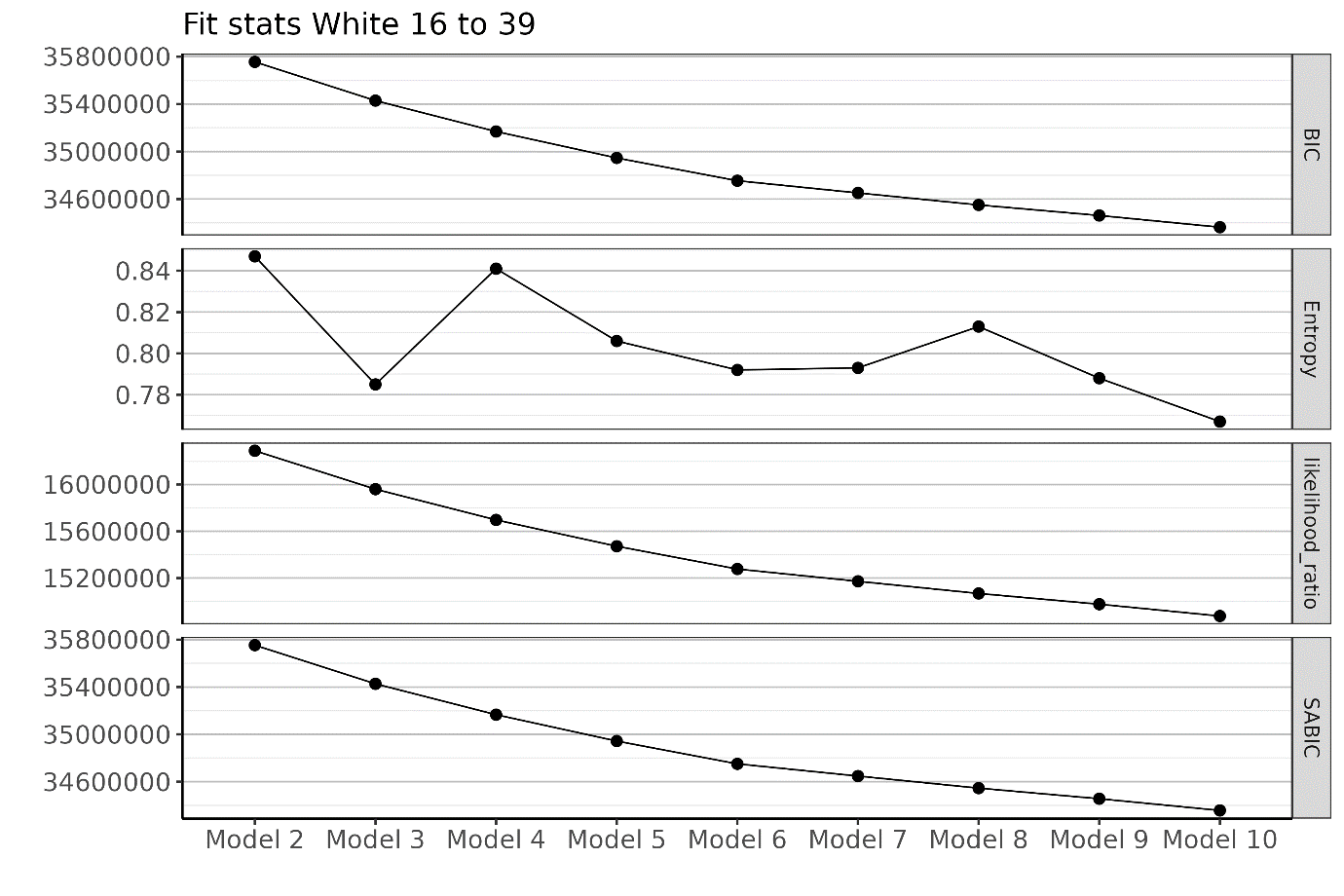

**B: South Asian**

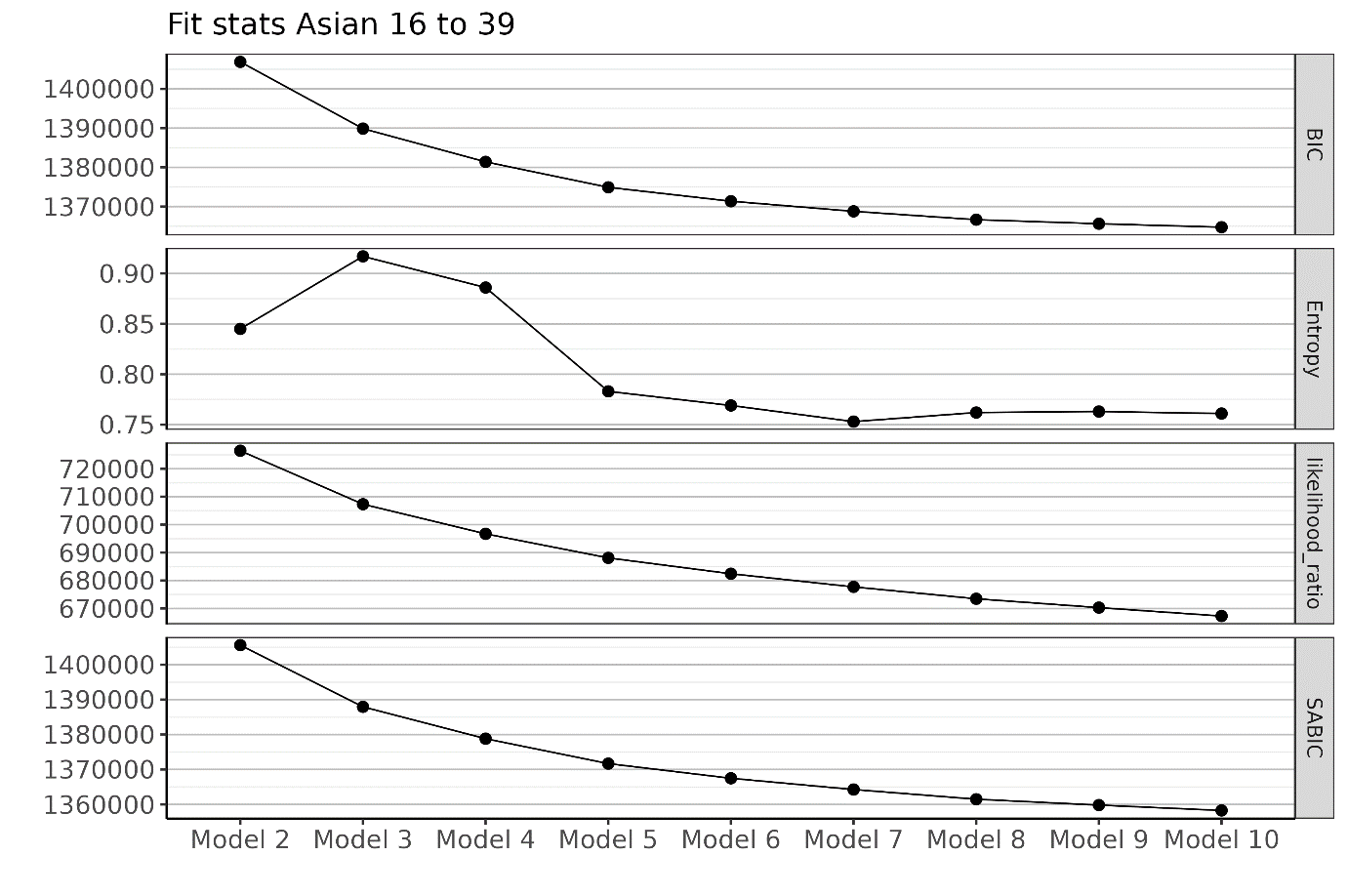

**C: Black or Black British**

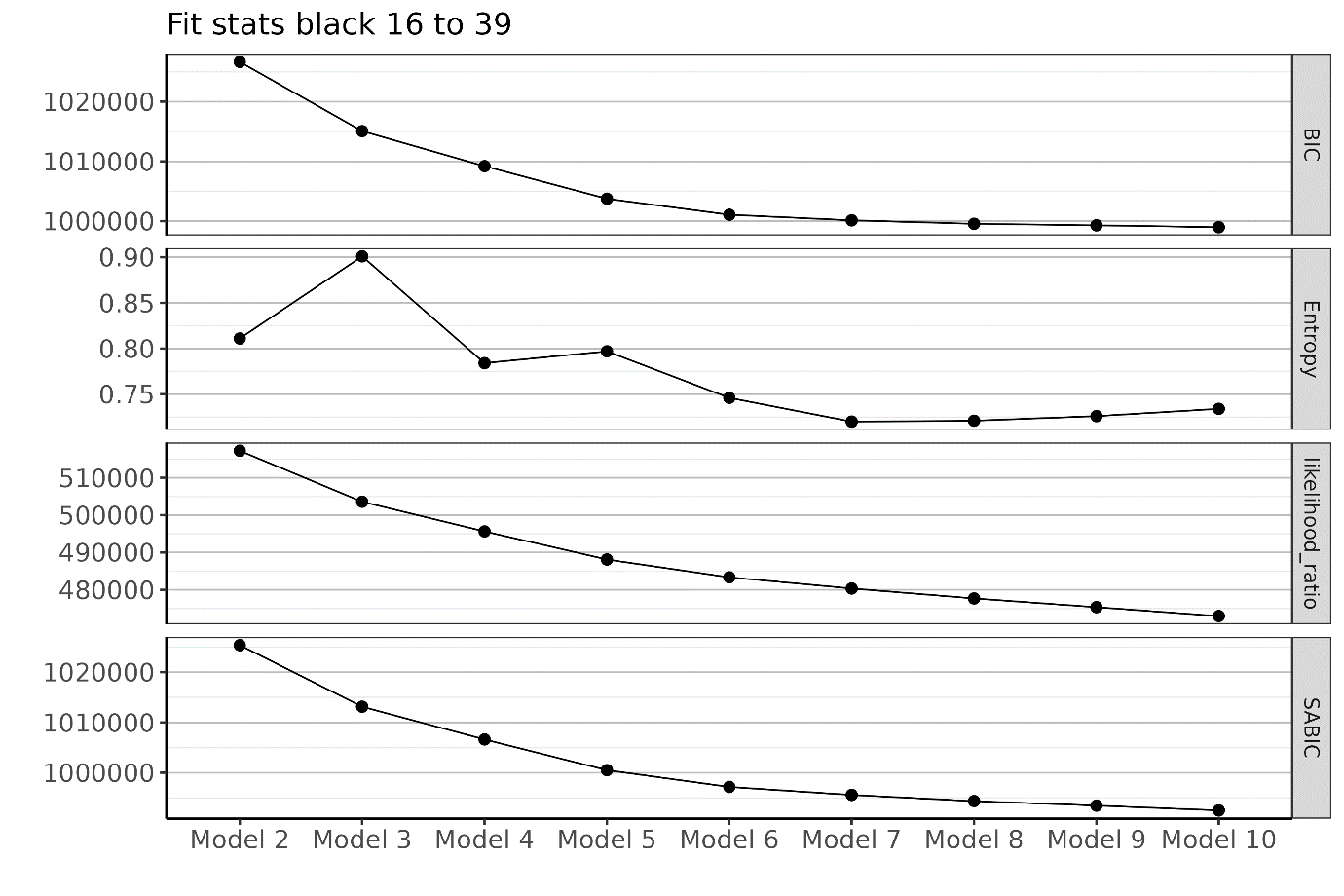

**Figure 1** - Fit statistics of the latent class models for the three ethnic groups.

**Table S3.** Fit statistics for the latent class models according to ethnic groups.

| **Model** | **log_likelihood** | **resid.df** | **BIC** | **AIC** | **Chisq** | **SABIC** | **likelihood_ratio** | **error_prior** | **error_post** | **Entropy** | **Group** |
| --- | --- | --- | --- | --- | --- | --- | --- | --- | --- | --- | --- |
| Model 2 | -701317.3213 | 33508 | 1406880.309 | 1403448.643 | 2.92E+45 | 1405587 | 726451.9576 | 0.487175668 | 0.075752801 | 0.845 | asian |
| Model 3 | -691750.5748 | 33304 | 1389874.865 | 1384723.15 | 6.47E+44 | 1387933 | 707318.4647 | 0.796990011 | 0.066392919 | 0.917 | asian |
| Model 4 | -686450.0011 | 33100 | 1381401.766 | 1374530.002 | 3.15E+42 | 1378812 | 696717.3172 | 0.970368649 | 0.110215816 | 0.886 | asian |
| Model 5 | -682140.9822 | 32896 | 1374911.778 | 1366319.964 | 2.08E+40 | 1371673 | 688099.2795 | 1.37247251 | 0.297731198 | 0.783 | asian |
| Model 6 | -679304.0053 | 32692 | 1371365.873 | 1361054.011 | 1.26E+40 | 1367479 | 682425.3257 | 1.592144656 | 0.368201414 | 0.769 | asian |
| Model 7 | -676955.3779 | 32488 | 1368796.667 | 1356764.756 | 1.37E+37 | 1364262 | 677728.0708 | 1.76007319 | 0.43419922 | 0.753 | asian |
| Model 8 | -674825.1655 | 32284 | 1366664.291 | 1352912.331 | 5.87E+36 | 1361481 | 673467.6461 | 1.884758204 | 0.447879329 | 0.762 | asian |
| Model 9 | -673244.9008 | 32080 | 1365631.811 | 1350159.802 | 4.78E+37 | 1359800 | 670307.1164 | 1.950600018 | 0.462931579 | 0.763 | asian |
| Model 10 | -671739.8294 | 31876 | 1364749.717 | 1347557.659 | 1.64E+36 | 1358270 | 667296.9738 | 2.042375455 | 0.488299208 | 0.761 | asian |
| Model 2 | -511263.327 | 25641 | 1026664.906 | 1023340.654 | 5.51E+47 | 1025371 | 517243.0573 | 0.455722844 | 0.08632568 | 0.811 | black |
| Model 3 | -504431.1784 | 25437 | 1015074.819 | 1010084.357 | 8.43E+46 | 1013133 | 503578.7601 | 0.742674141 | 0.073690574 | 0.901 | black |
| Model 4 | -500461.1686 | 25233 | 1009209.01 | 1002552.337 | 3.14E+46 | 1006619 | 495638.7405 | 1.147301567 | 0.247630766 | 0.784 | black |
| Model 5 | -496697.1553 | 25029 | 1003755.193 | 995432.3105 | 1.43E+45 | 1000517 | 488110.7137 | 1.266181127 | 0.256851823 | 0.797 | black |
| Model 6 | -494317.4525 | 24825 | 1001069.997 | 991080.9049 | 1.10E+44 | 997183.3 | 483351.3082 | 1.585033643 | 0.402742583 | 0.746 | black |
| Model 7 | -492812.2738 | 24621 | 1000133.85 | 988478.5475 | 8.31E+42 | 995598.9 | 480340.9507 | 1.803276012 | 0.50546652 | 0.72 | black |
| Model 8 | -491486.4095 | 24417 | 999556.3315 | 986234.8189 | 6.63E+44 | 994373.1 | 477689.2221 | 1.927614117 | 0.538056189 | 0.721 | black |
| Model 9 | -490320.697 | 24213 | 999299.1167 | 984311.394 | 2.41E+40 | 993467.5 | 475357.7972 | 1.952536796 | 0.535075245 | 0.726 | black |
| Model 10 | -489129.1085 | 24009 | 998990.1498 | 982336.2171 | 3.13E+40 | 992510.3 | 472974.6203 | 2.066843641 | 0.550229807 | 0.734 | black |
| Model 2 | -17875002.03 | 777497 | 35755551.88 | 35750822.06 | 2.15E+154 | 35754252 | 16289294.86 | 0.537737544 | 0.082073425 | 0.847 | white |
| Model 3 | -17710483.25 | 777292 | 35429295.02 | 35422194.51 | 8.42E+146 | 35427344 | 15960257.31 | 0.835281984 | 0.179459958 | 0.785 | white |
| Model 4 | -17578966.11 | 777087 | 35169041.43 | 35159570.22 | 3.54E+146 | 35166439 | 15697223.03 | 1.011667444 | 0.161118418 | 0.841 | white |
| Model 5 | -17466348.58 | 776882 | 34946587.06 | 34934745.16 | 4.70E+144 | 34943333 | 15471987.96 | 1.268809142 | 0.245637368 | 0.806 | white |
| Model 6 | -17368956.37 | 776677 | 34754583.34 | 34740370.74 | 5.02E+136 | 34750678 | 15277203.55 | 1.481724293 | 0.308854985 | 0.792 | white |
| Model 7 | -17316274.92 | 776472 | 34652001.14 | 34635417.85 | 4.07E+137 | 34647444 | 15171840.65 | 1.579234059 | 0.326786088 | 0.793 | white |
| Model 8 | -17264261.5 | 776267 | 34550755 | 34531801.01 | 4.33E+135 | 34545546 | 15067813.81 | 1.651448573 | 0.309527659 | 0.813 | white |
| Model 9 | -17218389.28 | 776062 | 34461791.24 | 34440466.56 | 7.58E+135 | 34455931 | 14976069.37 | 1.817802696 | 0.384675542 | 0.788 | white |
| Model 10 | -17167755.81 | 775857 | 34363305 | 34339609.63 | 3.84E+134 | 34356793 | 14874802.43 | 2.014486824 | 0.469591847 | 0.767 | white |

4. **Supplementary table and figures**

| **Table S4.** The 21 common LTC to the three ethnic groups and their highest prevalence across all clusters and the population prevalence. | | | | | | | | | |
| --- | --- | --- | --- | --- | --- | --- | --- | --- | --- |
|  | **White** | | | **South Asian** | | | **Black or Black British** | | |
| **Long-term condition** | **% population** | **% cluster** | **Cluster** | **% population** | **% cluster** | **Cluster** | **% population** | **% cluster** | **Cluster** |
| Allergic and chronic rhinitis | 21 | 31 | 2 | 28 | 39 | 2 | 29 | 38 | 2 |
| Anxiety and phobia | 34 | 53 | 2 | 17 | 32 | 2 | 16 | 29 | 2 |
| Asthma | 25 | 32 | 2 | 20 | 32 | 2 | 18 | 30 | 2 |
| Constipation | 18 | 37 | 3 | 23 | 43 | 2 | 21 | 39 | 2 |
| Depression | 49 | 72 | 2 | 26 | 49 | 2 | 29 | 49 | 2 |
| Dermatitis (atopic/contact/other/unspecified) | 30 | 40 | 2 | 34 | 48 | 2 | 25 | 35 | 2 |
| Ddysmenorrhoea | 8 | 11 | 4 | 8 | 10 | 3 | 10 | 15 | 3 |
| Enthesopathies and synovial disorders | 26 | 47 | 2 | 20 | 48 | 2 | 16 | 37 | 2 |
| Female infertility | 8 | 100 | 4 | 13 | 100 | 3 | 12 | 100 | 3 |
| Gastritis and duodenitis | 12 | 30 | 3 | 13 | 34 | 2 | 10 | 26 | 2 |
| Gastro-oesophageal reflux disease | 17 | 36 | 2 | 18 | 42 | 2 | 13 | 30 | 2 |
| Hypertension | 18 | 70 | 3 | 16 | 55 | 2 | 21 | 60 | 2 |
| Male infertility | 7 | 88 | 4 | 12 | 88 | 3 | 10 | 86 | 3 |
| Menorrhagia and polymenorrhoea | 17 | 40 | 2 | 17 | 31 | 2 | 21 | 35 | 2 |
| Migraine | 16 | 30 | 2 | 14 | 24 | 2 | 13 | 19 | 2 |
| Obesity | 18 | 32 | 3 | 16 | 31 | 2 | 22 | 38 | 2 |
| Osteoarthritis | 13 | 41 | 3 | 8 | 31 | 2 | 7 | 28 | 2 |
| Other anaemia | 10 | 26 | 3 | 22 | 41 | 2 | 21 | 37 | 2 |
| Painful conditions | 22 | 57 | 3 | 21 | 62 | 2 | 17 | 52 | 2 |
| Post-traumatic stress and stress-related disorders | 22 | 41 | 2 | 16 | 27 | 2 | 18 | 31 | 2 |
| Type 2 diabetes | 6 | 26 | 3 | 11 | 36 | 2 | 7 | 25 | 2 |

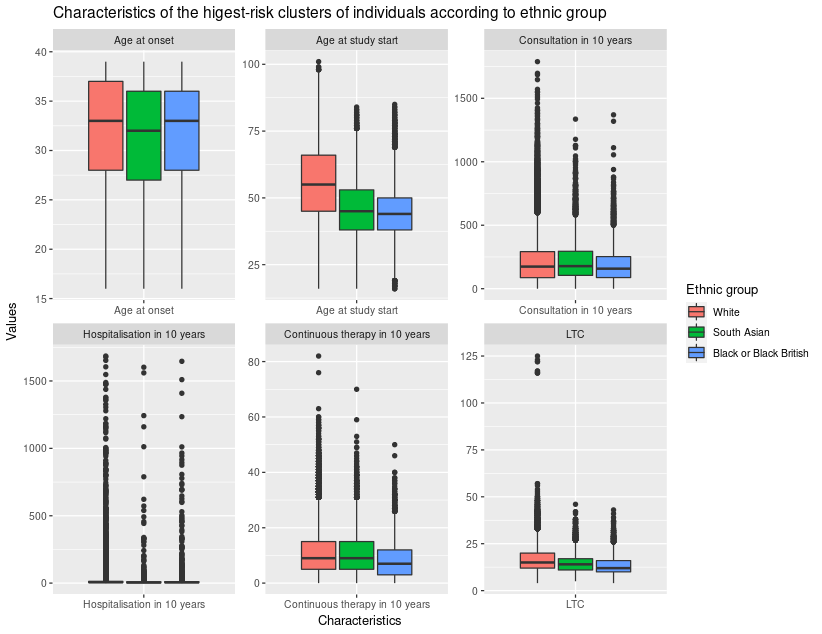

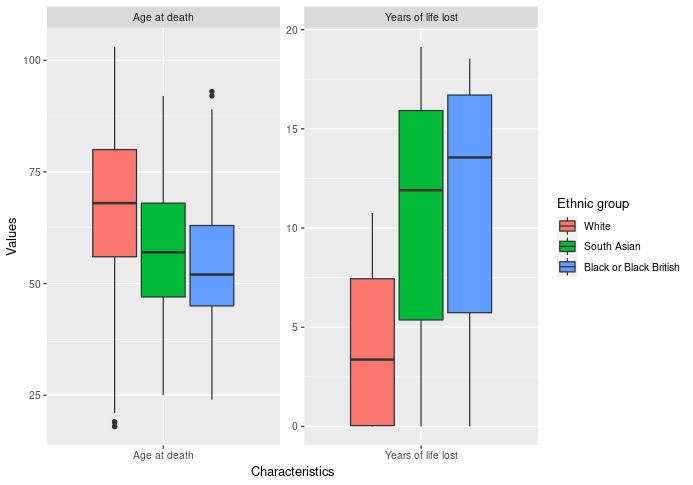

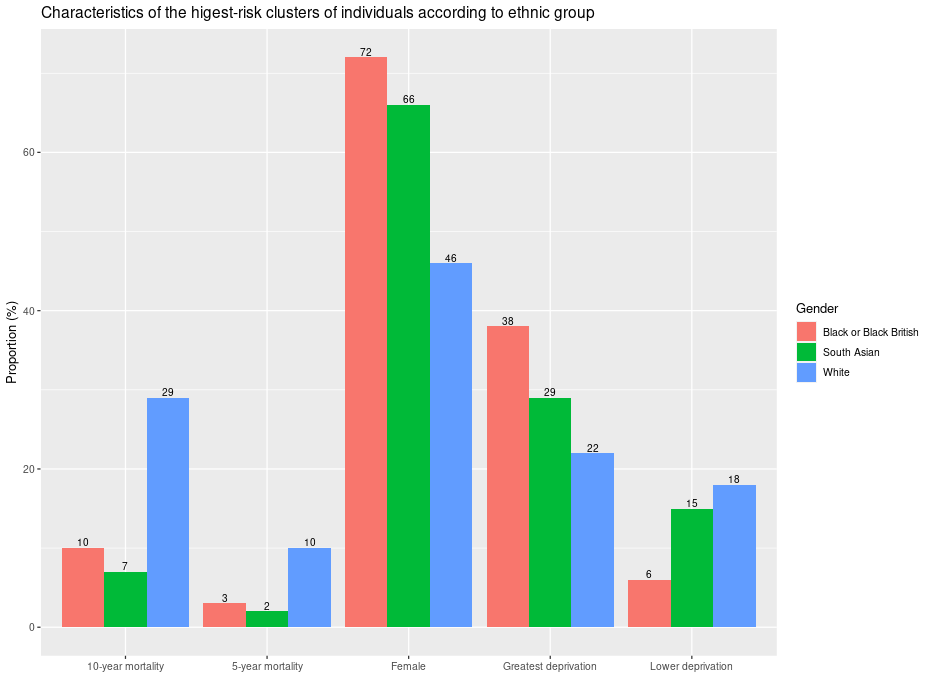

**Figure 3** - Characteristics of the highest-risk clusters of individuals according to ethnic groups
